# Machine Learning Reveals the Contribution of Rare Genetic Variants and Enhances Risk Prediction for Coronary Artery Disease in the Japanese Population

**DOI:** 10.1101/2024.08.13.24311909

**Authors:** Hirotaka Ieki, Kaoru Ito, Sai Zhang, Satoshi Koyama, Martin Kjellberg, Hiroki Yoshida, Ryo Kurosawa, Hiroshi Matsunaga, Kazuo Miyazawa, Nobuyuki Enzan, Changhoon Kim, Jeong-Sun Seo, Koichiro Higasa, Kouichi Ozaki, Yoshihiro Onouchi, The Biobank Japan Project, Koichi Matsuda, Yoichiro Kamatani, Chikashi Terao, Fumihiko Matsuda, Michael Snyder, Issei Komuro

## Abstract

Genome-wide association studies (GWASs) have advanced our understanding of coronary artery disease (CAD) genetics and enabled the development of polygenic risk scores (PRSs) for estimating genetic risk based on common variant burden. However, GWASs have limitations in analyzing rare variants due to insufficient statistical power, thereby constraining PRS performance. Here, we conducted whole genome sequencing of 1,752 Japanese CAD patients and 3,019 controls, applying a machine learning-based rare variant analytic framework. This approach identified 59 CAD-related genes, including known causal genes like *LDLR* and those not previously captured by GWASs. A rare variant-based risk score (RVS) derived from the framework significantly predicted CAD cases and cardiovascular mortality in an independent cohort. Notably, combining the RVS with traditional PRS improved CAD prediction compared to PRS alone (area under the curve, 0.66 vs 0.61; p=0.007). Our analyses reinforce the value of incorporating rare variant information, highlighting the potential for more comprehensive genetic assessment.

## Introduction

Despite advancements in treatments and medications, coronary artery disease (CAD), encompassing conditions such as angina pectoris and myocardial infarction (MI), remains a leading cause of death worldwide ^1,2^. CAD etiology is complex, involving a multifaceted interplay between genetic predisposition and environmental determinants. Lifestyle factors including diet, smoking, and physical activity are well-established contributors to the onset and progression of CAD ^3,4^. Additionally, conditions such as elevated low-density lipoprotein (LDL) cholesterol, hypertension, and glucose intolerance further exacerbate the risk profile ^5^. The importance of genetic predisposition is also underscored by a European twin study, which estimated that genetic factors contributed to over 50% of CAD development ^6,7^. Therefore, understanding the genetic underpinnings of CAD and accurately estimating an individual’s lifetime genetic risk are crucial for effective prevention and management strategies.

To date, genome-wide association studies (GWASs) and their meta-analyses have identified more than 300 loci associated with CAD ^8–12^. Polygenic risk scores (PRSs) derived from GWAS summary statistics have enabled the estimation of individual-level CAD risk ^13,14^. However, despite these significant advancements, the heritability of CAD explained by GWASs remains lower than anticipated. This gap may be partly attributed to the primary focus of GWAS on low frequency to common variants, while rare variants are often underrepresented in these analyses ^5,15^. Rare variants often have a large effect size on diseases and phenotypes, making them a promising target for drug development ^16^. Incorporating rare variants into genetic risk scores could significantly enhance the accuracy of CAD prediction. Despite this potential, previous GWASs and aggregated rare variant association analyses have struggled even in large-scale sequencing studies, identifying only a few genes at exome-wide significance per trait ^17,18^. Furthermore, calculating a genetic risk score based on rare variants is challenging because gene-level effect sizes are not estimated by conventional gene-based analysis methods.

Recently, advancements in machine learning have led to the development of novel methods for genetic analysis, one of which is the HEAL (Hierarchical Estimate from Agnostic Learning) method, a machine learning-based framework for comprehensive rare variant analysis. This approach has been successful in identifying disease-associated genes and creating genetic risk scores in patients with abdominal aortic aneurysm ^19^. In the current study, we conducted whole genome sequencing (WGS) of Japanese CAD patients and applied a modified version of the HEAL framework tailored for CAD to analyze rare variants and systematically prioritize disease-associated genes. Furthermore, we developed a rare variant-based genetic risk score (RVS) using this framework and validated the performance with an independent cohort. We then explored the relationship between the RVS and GWAS-based PRS to elucidate the characteristics of rare variants in CAD, bridging the gap in our understanding of CAD genetics by incorporating rare variant information, potentially uncovering novel insights into disease mechanisms and improving risk prediction models.

## Results

### Whole genome sequencing of CAD samples in the Japanese population

The overview and the design of our study are shown in **Figure 1**. We performed WGS on the discovery cohort comprising 1,765 Japanese CAD patients and 3,148 controls. In order to enhance the genetic discovery power ^20^, we prioritized patients with early-onset MI, a severe form of CAD, from the BioBank Japan (BBJ) cohort. The average age of MI onset in these patients was 47.4 ± 4.1 years, indicating a relatively young population with a severe disease phenotype. After quality control of the WGS data, we retained 4,771 individuals (1752 cases and 3019 controls) with 51,717,580 genetic variants. For the validation WGS cohort, we included 200 CAD cases and 824 control samples with 25,531,471 variants (**Table S1 and S2**). Demographic features in each cohort are summarized in **Table 1**. We then used the quality-controlled data for further analyses including single variant association tests to identify individual variants associated with CAD, a conventional gene-based association test to examine the cumulative effect of variants within specific genes, and a machine learning-based framework to uncover the potential contribution of rare variants (**Figure S1**).

**Figure 1.**
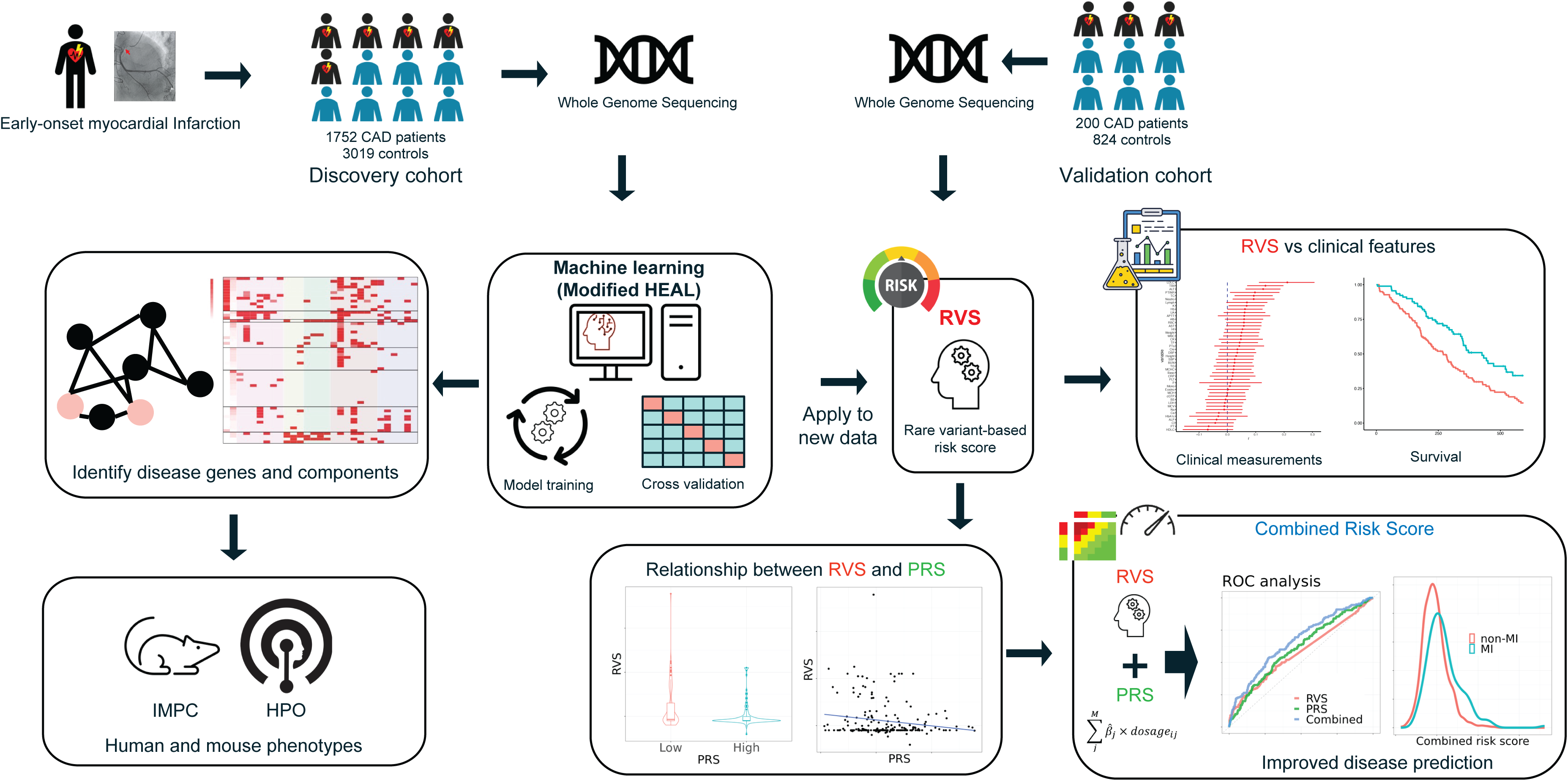
Overview of the current study. We studied the genetic factors of coronary artery disease (CAD) combining whole-genome sequencing data and a machine learning-based framework named the modified HEAL method in patients with MI, one of the most severe forms of CAD, and controls. We sequenced the whole genomes of Japanese CAD patients and controls and applied the modified HEAL method framework. The framework was based on a sparse modeling devised to distinguish diseased individuals from controls. After the hyperparameter tuning and training of the model by the cross-validation method, the model outputted a list of genes related to CAD, which were subsequently analyzed by a clustering-based method and mapped on the protein-protein interaction network to reveal the CAD-associated modules. The function of the identified genes was also confirmed by the human phenotype and knockout mouse phenotype databases. The learned (optimized) machine learning model was applied to derive rare variant-based genetic risk scores (RVS) to predict CAD outcomes in an independent validation cohort. We also tested the relationship of the RVS with clinical features and common variant-based polygenic risk score (PRS). RVS was combined with PRS to improve the prediction performance of CAD disease status in the independent validation cohort. BBJ, BioBank Japan; MI, myocardial infarction; CRS, combined risk score

**Table 1.** Demographic features of participants.

| Data | Disease status | Total | Males |  | Age (years) |  | BMI (kg/m<sup>2</sup>) |  | Age at MI onset (years) |  |
| --- | --- | --- | --- | --- | --- | --- | --- | --- | --- | --- |
|  |  | N | N | % | Mean | SD | Mean | SD | Mean | SD |
| Discovery cohort | Case | 1,752 | 1,617 | 92.29 | 60.1 | 13.7 | 25.0 | 3.4 | 47.4 | 4.1 |
|  | Control | 3,019 | 1,205 | 39.91 | 55.3 | 8.0 | 23.9 | 4.0 | - | - |
| Validation cohort | Case | 200 | 183 | 91.50 | 43.8 | 9.8 | 26.5 | 4.3 | 36.0 | 3.9 |
|  | Control | 824 | 420 | 50.97 | 49.3 | 13.0 | 22.9 | 3.8 | - | - |

**SD, standard deviation; BMI, body mass index; MI, myocardial infarction**

We first conducted a single variant association test in the discovery cohort using a logistic regression model implemented in PLINK software with covariates of age, sex and top ten ancestry principal components (PCs). The genomic inflation factor λ*_GC_* was calculated to be 1.03, indicating minimal inflation of test statistics and suggesting that the quality control applied to the samples was adequate (**Figure S2**). This initial single variant association analysis did not identify any genetic loci that reached a genome-wide significance threshold of P = 5 * 10^-8^. A subsequent analysis was performed using SAIGE software designed to handle both common and rare variants, adjusted for age, sex and top ten ancestry PCs. This analysis revealed two previously reported loci on chromosome 12 that reached a genome-wide significance threshold (rs7977233; p=1.47 * 10^-8^, rs3782886; p=1.47 * 10^-8^, respectively, (**Figure S3 and Table S3**))^10,11^. However, these were both common variants, emphasizing the difficulty in analyzing rare variants using current GWAS approaches.

To increase the detection power of rare variant associations, gene-based tests are often used, in which variants are aggregated and analyzed together for each gene. This approach allows for the analysis of rare variants that are underpowered in single variant association tests due to their low frequency. It also increases detection power by reducing the multiple testing burden. Thus, we conducted a gene-based rare variant aggregated association analysis using the sequential kernel association test-optimal (SKAT-O). While no genomic inflation was observed (λ = 0.939) (**Figure S4**), the *LDLR* gene surpassed a suggestive threshold (p = 2.3×10^-5^). However, no genes reached the gene-wide significance threshold of p = 2.5×10^-6^ (**Figure S4 and Table S4**). This result also highlighted the challenges of analyzing rare variants in genetic association studies due to insufficient statistical power with a limited sample size.

### The machine learning-based framework prioritizes disease-associated genes and reveals molecular networks

We next conducted a machine learning-based rare variant analysis using a modified HEAL ^19^. In this framework, we first quantified the mutation burden for each gene in each participant defined by the cumulative effects of deleterious nonsynonymous variants within the gene. We then trained a penalized logistic regression model to predict disease status based on these mutation burden scores. The model was trained to identify a minimal set of most distinguishing features (genes) for CAD, while also optimizing parameters for accurate disease prediction. Through robust cross-validation (**Figure S5**), we successfully prioritized fifty-nine candidate genes associated with CAD development (**Table S5, S6 and Figure S6)**.

To investigate the functions of the fifty-nine HEAL_CAD_ genes, we assessed constraint scores and checked for overlaps with neighboring genes identified in previous GWASs on CAD and its risk factors. Using the Genehancer database ^21^, which provides information on genome-wide enhancers and their target genes, we identified prioritized genes that overlapped with the target genes of enhancers found significant in previous GWASs. We also referenced the International Mouse Phenotyping Consortium (IMPC) ^22^ database to investigate the phenotypes associated with a gene knockout (KO) in mice and conducted gene set enrichment analysis to identify functional clusters among the HEAL_CAD_ genes. The genes were subsequently categorized into eight distinct clusters based on the hierarchical clustering of their functional annotations (**Figure 2A, 2B and Table S7**).

**Figure 2.**
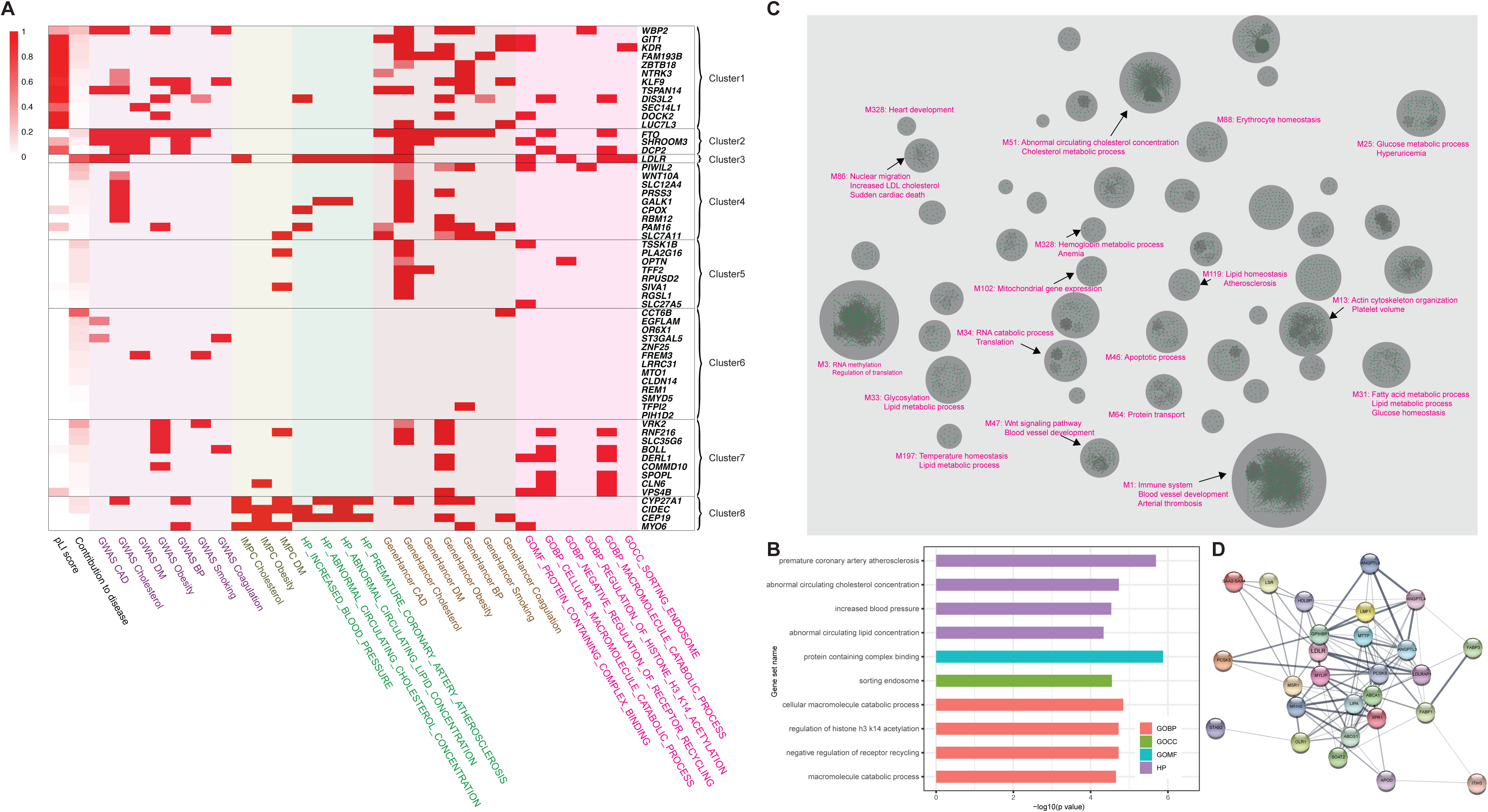
Functional analysis of HEAL_CAD_ genes. (A) Fifty-nine genes identified by the machine learning-based framework were annotated using six different criteria; 1) The constraint score (pLI) from the gnomAD database 2) Overlap with GWAS on CAD and its risk factor (lipids, diabetes, obesity, blood pressure, coagulation, smoking) phenotypes, 3) Overlap with the genes in which GWAS-significant variants act as enhancers, 4) Knock-out mouse phenotype with blood pressure, diabetes, and lipid traits, 5) Human phenotype ontology and 6) Gene ontology. Then the fifty-nine genes were grouped into eight clusters by hierarchical clustering based on functional annotations. For GWAS and Genehancer, red indicates a significant association and light red denotes suggestive significance. (B) Gene ontology (GO) and human phenotype ontology (HPO) term enrichment analysis. The GO and HPO annotation results were based on 59 genes. Gene ontology categories included molecular function, cellular components and biological process. GO and HPO categories for each function were sorted by decreasing order of evidence based on the GO enrichment test P-value. Only the significant categories after multiple test corrections are shown. (C) The forty-six modules were identified in the protein-protein-interaction network using diffusion component analysis seeded by the 59 HEAL_CAD_ genes. (D) Visualization of the module 119 network of the protein-protein interactions. The module included important genes involved in cholesterol metabolism, including *LDLR*, *PCSK9*, *ANGPTL3*, *ANGPTL4*, and *LIPA*. GWAS, genome-wide association study; CAD, coronary artery disease; DM, diabetes mellitus; BP, blood pressure; IMPC, International Mouse Phenotyping Consortium; HP, human phenotype; GOMF, gene ontology molecular function; GOBP, gene ontology biological pathway; GOCC, gene ontology cellular component.

Among these clusters, cluster 3 notably included the *LDLR* gene, which exhibited the strongest contribution to CAD. *LDLR* is a well-established causal gene for familial hypercholesterolemia ^23^ and has been consistently associated with CAD in previous GWASs and genome sequencing studies ^9,24,25^, supporting the validity of our machine learning-based framework. In the IMPC database, *LDLR* KO mice showed increased circulating cholesterol levels ^26^, a known risk factor for CAD. Cluster 7 contained genes related to obesity and metabolic processes, such as the *RNF216* locus, which is associated with body mass index (BMI) ^27^ and increased glucose levels in KO mice ^22^. Additionally, the *VRK2* locus has been reported to be associated with BMI ^28^, smoking behavior and alcohol use ^29^, indicating its broader impact on metabolic health. Cluster 2 comprised genes identified by previous GWAS on phenotypes such as blood pressure, diabetes, and cholesterol levels. The *FTO* gene within this cluster was highlighted for its strong association with obesity ^30,31^ and related phenotypes linked to BMI ^32^, LDL cholesterol ^33^, blood pressure ^34^, and CAD ^35^. Cluster 8 encompassed genes associated with cholesterol levels, obesity and blood pressure in GWAS and GeneHancer categories, with phenotypic evidence in human and KO mice. For instance, the *CYP27A1* locus is associated with diastolic blood pressure ^36^ and triglyceride levels ^37^ and has connections to cholesterol levels and premature CAD according to human phenotype ontology ^38^.

To further determine the functions of the fifty-nine genes, we mapped them onto the human protein-protein interaction (PPI) network followed by identifying proteins that were tightly clustered with these HEAL_CAD_ genes as topological modules ^19^. We identified 46 tightly clustered topological modules encompassing the HEAL_CAD_ genes. Gene ontology analysis confirmed the functional coherence of the proteins within each module, revealing significant enrichment for specific biological processes. For instance, module M119 was significantly enriched for lipid homeostasis with a false discovery rate (FDR) of 2.53*10^-22^, suggesting a critical role in regulating lipid levels (**Figure 2C and Table S8**). These modules included pathways known as CAD risk factors, such as lipid and glucose metabolism (M25, M31, M51, M86, M119). Notably, M119 included lipid metabolism-related genes such as *LDLR*, *PCSK9*, *LIPA,* and *ANGPTL3* (**Figure 2D**), which are well-known targets for medications treating dyslipidemia and CAD ^39^ ^40^. Other modules were associated with different biological processes, including platelet volume (e.g., M13), immune system function (M1), blood vessel and heart development (e.g., M47, M328), and RNA metabolism and translation processes (e.g., M3, M34). While recent studies have indicated the contribution of common variants identified by CAD-GWAS to the disease through various pathways such as plaque formation, inflammation, transcriptional regulation, and angiogenesis ^41^, our findings suggest that diverse biological processes are also implicated in CAD, even in the context of rare variants. This underscores the complexity of CAD pathogenesis, involving a wide array of biological pathways and molecular mechanisms.

### Rare variant risk-based risk score and its clinical impact

In conjunction with the prioritization of disease-related genes, the modified HEAL enabled us to develop a prediction model for CAD based on genetic information. Using the optimized machine learning model, we computed a rare variant-based risk score (RVS) for each individual. The RVS demonstrated a significant predictive capability for CAD, with an area under the receiver operating characteristics curve (AUROC) of 0.574, as validated through a nested cross-validation approach in the discovery cohort. When applied to an independent validation cohort, the RVS also identified CAD cases with an AUROC of 0.581 (p = 0.002), indicating its ability to discriminate CAD cases.

To further understand the characteristics of RVS in terms of clinical aspects, we explored the association of RVS with clinically relevant parameters. The RVS showed significant correlations with several key clinical measurements, including low-density-lipoprotein cholesterol (LDLC), total bilirubin (TBil), alanine aminotransferase (ALT), prothrombin time (PT-INR), total cholesterol levels, neutrophil count, and potassium levels (**Figure 3A and Table S9**). These correlations are noteworthy since elevated cholesterol levels and coagulation abnormalities are established risk factors for CAD ^42–44^. Moreover, alterations of total bilirubin and AST were also reported to be associated with cardiovascular risk ^45,46^, reinforcing the clinical relevance of the RVS in the context of CAD.

**Figure 3.**
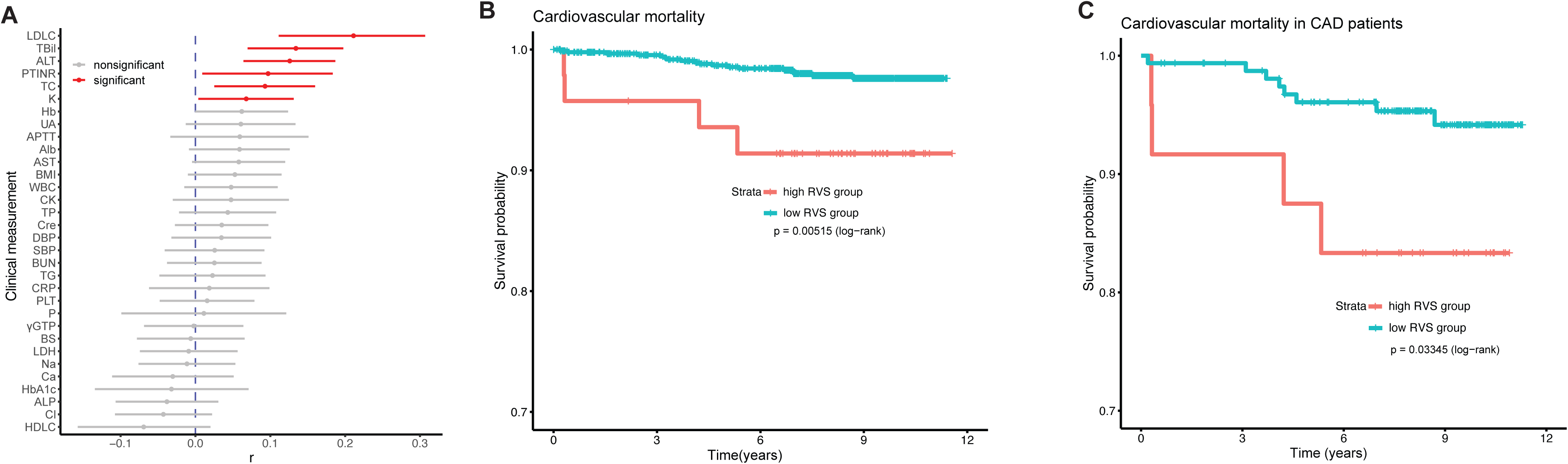
Rare variant risk score (RVS) and its clinical impact. (A) Correlation between RVS and continuous clinical indices. Data are presented as Pearson’s correlation coefficients and their 95% confidence intervals (CIs). Exact P values are shown in Table S9. (B) Kaplan-Meier curves for cardiovascular mortality among total participants stratified into two groups based on RVS. Participants with high RVS died significantly earlier than those with low RVS. (C) Kaplan-Meier curves for cardiovascular mortality among CAD patients (n=200) stratified into two groups based on RVS. CAD patients with high RVS (top 5%) showed significantly worse cardiovascular prognosis. LDLC, low-density lipoprotein cholesterol; Tbil, total bilirubin; ALT, alanine aminotransferase; PTINR, prothrombin time international normalized ratio; TC, total cholesterol; K, potassium; Hb, hemoglobin; UA, uric acid; APTT, activated partial thromboplastin time; Alb, albumin; RBC, red blood cell; AST, aspartate aminotransferase; WBC, white blood cell; CK, creatine kinase; TP, total protein; Cre, creatinine; DBP, diastolic blood pressure; SBP, systolic blood pressure; BUN, blood urea nitrogen; TG, triglycerides; CRP, C-reactive protein; PLT, platelet; P, Phosphorus; _γ_GTP, gamma-glutamyl transpeptidase; BS, blood sugar; LDH, Lactate dehydrogenase.

We extended our analysis to assess the impact of the RVS on long-term cardiovascular mortality. In the validation cohort, a higher RVS was significantly associated with increased cardiovascular mortality (P = 0.01, log-rank test) (**Figure 3B)**. When exclusively analyzing CAD patients, those with higher RVS also exhibited a significantly worse cardiovascular mortality rate (p = 0.03, log-rank test) (**Figure 3C**). These findings suggest that RVS not only predicts CAD occurrence but also correlates with the disease severity and its long-term prognosis, highlighting its potential clinical utility in risk stratification and prognosis estimation for CAD patients.

### The integration of RVS and PRS improves the performance of the genomic risk score

Many GWASs have been conducted for CAD, leading to the development of PRS that primarily comprise common variants to predict the risk of CAD. Multiple studies have reported that PRS can serve as an important indicator for predicting and assessing the severity of CAD. Whereas these scores typically focus on common variants and do not account for rare variants, which can also significantly contribute to disease risk, our RVS encompasses rare variants not included in PRS. Thus, to compare the properties between RVS and PRS, we first calculated individual PRS based on CAD-GWAS ^11^ in the validation cohort. The PRS also significantly predicted CAD with an AUROC of 0.61 (p = 0.001; 95% confidence interval (C.I.), 0.565-0.653). Interestingly, there was no significant correlation between PRS and RVS (r = -0.01, p = 0.73) (**Figure 4A**), indicating that RVS provides a different genomic perspective on CAD risk.

**Figure 4.**
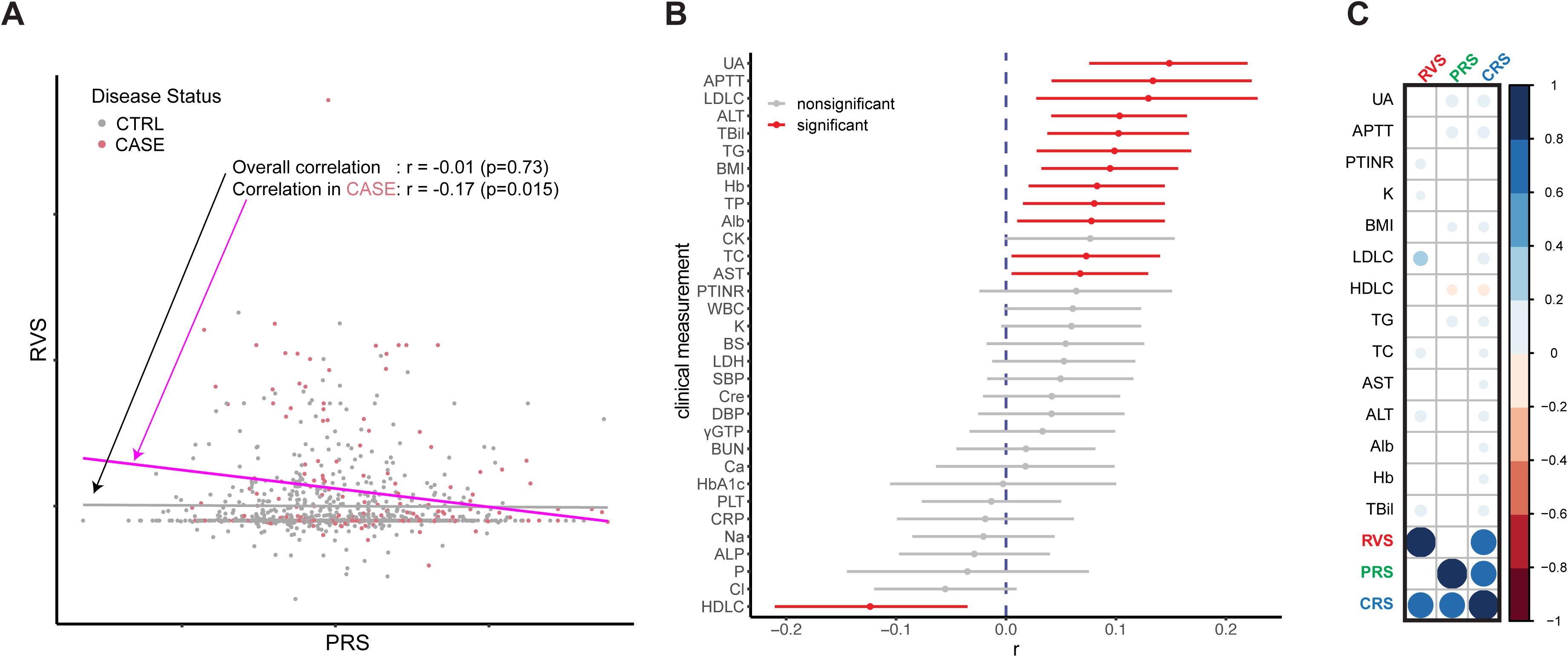
The Relationship between RVS, PRS, CRS, and clinical indices. (A) A scatter plot illustrating the relationship between RVS and PRS, with cases (red) and controls (gray) color-coded. The overall (gray) and case-only (pink) regression lines and correlation coefficients are shown. A significant negative correlation was observed in the CAD cases. (B) Correlation between combined risk score (CRS), defined by the average of RVS and PRS, and continuous clinical indices. Data are presented as Pearson’s correlation coefficients and their 95% CIs. Exact P values are shown in Table S11. (C) Correlation between clinical measurements and different genetic risk scores (RVS, PRS and CRS). Only significant correlations are displayed with a circle. Blue, positive correlation; red, negative correlation. Larger circles correspond to a stronger correlation. LDLC, low density lipoprotein cholesterol; Tbil, total bilirubin; ALT, alanine aminotransferase; PTINR, prothrombin time international normalized ratio; TC, total cholesterol; K, potassium; Hb, hemoglobin; UA, uric acid; APTT, activated partial thromboplastin time; Alb, albumin; RBC, red blood cell; AST, aspartate aminotransferase; WBC, white blood cell; CK, creatine kinase; TP, total protein; Cre, creatinine; DBP, diastolic blood pressure; SBP, systolic blood pressure; BUN, blood urea nitrogen; TG, triglycerides; CRP, C-reactive protein; PLT, platelet; P, Phosphorus; _γ_GTP, gamma-glutamyl transpeptidase; BS, blood sugar; LDH, Lactate dehydrogenase

When examining CAD cases specifically, RVS showed a negative correlation with PRS (r = -0.17, p = 0.015) (**Figure 4A**). Additionally, PRS was associated with different clinical measurements compared to RVS, such as triglycerides, uric acid, body mass index (BMI), and activated partial thromboplastin time (APTT) and it was negatively associated with HDL cholesterol (HDLC), which is considered protective against CAD (**Figure 3A, Figure S7 and Table S10**) ^47^. These data support the notion that PRS and RVS may have complementary rather than redundant roles in predicting CAD, as they were associated with different clinical parameters and did not show a positive correlation.

Given these distinct properties, we integrated PRS and RVS to develop a combined risk score (CRS) aiming at enhancement of the performance of the framework in predicting CAD. The CRS showed positive correlations with several clinical measures, including serum urinary acid, coagulation functions, LDLC, and triglycerides (TG), while negatively correlating with HDLC levels (**Figure 4B and Table S11**). Focusing on lipid metrics, CRS demonstrated correlations with LDLC, TC, TG, and HDLC, suggesting that it combines the unique predictive elements of both RVS and PRS (**Figure 4C**). Finally, we evaluated the predictive performance of CRS and observed a significant improvement in CAD prediction compared to PRS alone in the validation cohort (AUROC 0.66 vs 0.61, p=0.007; Pseudo R^2^ 0.093 vs 0.040, p = 0.0018; AUPRC 0.35 vs 0.29, p = 0.0154) (**Figure 5 and Table S12**). These results suggest that RVS can complement PRS and that incorporating rare variant information as an RVS into PRS significantly enhances the ability to predict CAD, thereby addressing some of the unexplained heritability in the disease.

**Figure 5.**
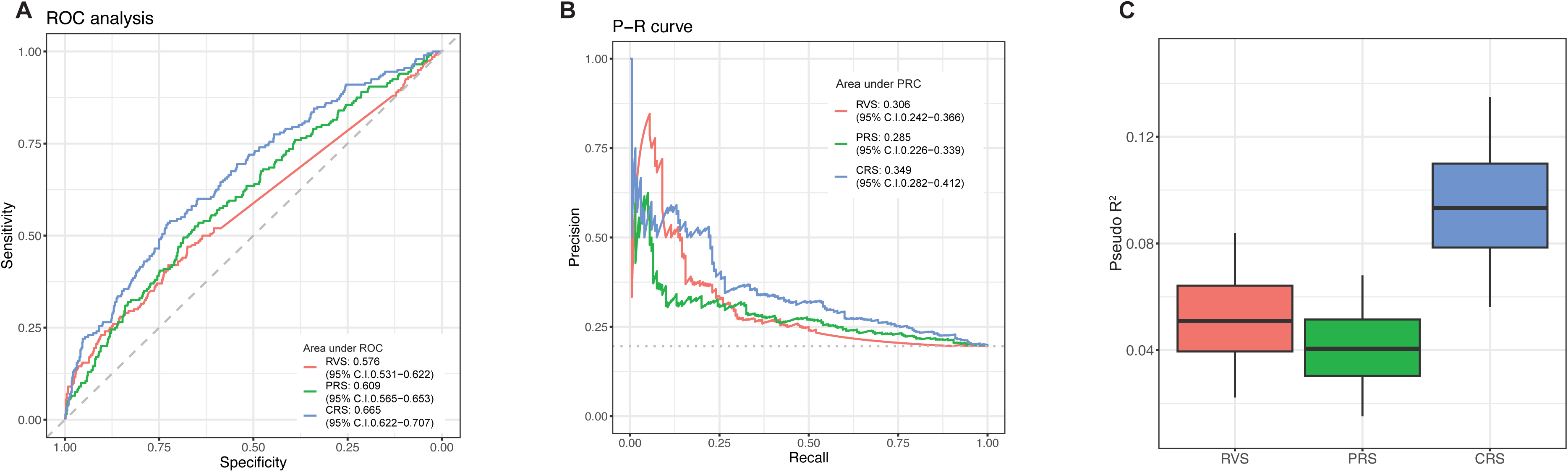
The combined RVS and PRS risk score improved CAD prediction. (A) Receiver operating characteristic (ROC) curve for RVS, PRS and CRS (Combined Risk Score). The curve plots the true positive rate (sensitivity) against the false positive rate (1-specificity) for different threshold values of the predictive score. The area under the curve (AUC) is indicated, representing the score’s accuracy in predicting the outcome. The dotted line represents a reference line of no discrimination (AUC = 0.5). Points on the curve closer to the top-left corner indicate higher diagnostic accuracy. (B) Precision-recall curve (PRC) for RVS, PRS and CRS. The curve shows the trade-off between precision (positive predictive value) and recall (sensitivity) at various threshold levels. The confidence interval for the area under the PRC was estimated from the 20,000 times bootstrap replication method. (C) Boxplot of Pseudo R^2^ for CAD prediction performance. This box plot displays the pseudo-R^2^ values comparing the CAD prediction performance of RVS, PRS and CRS. The distribution of pseudo-R^2^ was estimated from 20,000 times bootstrapping. The box plot center line represents the median, the bounds represent the first and third quartile, and the whiskers reach to 1.5 times the interquartile range.

## Discussion

In this study, we developed a machine learning-based analytical framework to investigate the genetics of CAD pathogenesis with a focus on rare variants. We leveraged this framework together with whole-genome sequencing (WGS) data from the Japanese population to enhance our understanding of the complex CAD genetic architecture. Our findings indicated that the modified HEAL, a machine learning-based framework, effectively prioritized genes associated with CAD, including the well-established *LDLR* gene, while also uncovering intricate molecular networks involved in the disease. The rare variant-based risk score (RVS) generated through this framework demonstrated significant predictive power for CAD and long-term cardiovascular mortality Furthermore, the RVS showed different characteristics from conventional common variant-based PRS, and combining the rare variant-based RVS with the PRS substantially improved CAD prediction.

Identifying disease-associated rare variants remains a significant challenge, not only in single variant association analyses but also in aggregated rare variant association analyses ^48,49^. While some studies have adopted a targeted resequencing approach by selecting specific genes based on prior knowledge ^25,50^; previous attempts at genome-wide or exome-wide analyses have often suffered from insufficient statistical power, leading to limited success in identifying previously uncharacterized genes associated with complex traits like CAD ^20^. Also in this study, the single variant association analysis and the gene-based rare variant association analysis failed to reveal genome-wide significant rare variants linked to CAD. Even in previous studies involving more than 450,000 exome sequencing data from the UK biobank, only a single gene, *LDLR*, reached a significance level in the gene-based test for CAD ^17^. These persistent challenges highlight the difficulties in rare variant analyses.

To address these challenges, we utilized a machine learning-based framework to analyze rare variants, building on the HEAL model in a prior study, where Li et al. successfully uncovered the genetic architecture of rare variants in abdominal aortic aneurysm ^19^. We adapted and optimized the model for CAD patients, marking the first application of the technique in this disease context. Unlike the previous HEAL model that focused only on missense single nucleotide variants (SNVs), our approach casts a wider net as it incorporates insertion, deletion and putative loss-of-function (pLOF) variants. This comprehensive inclusion of variant types allows for a more holistic examination of the genetic landscape underlying CAD, potentially capturing a broader spectrum of disease-associated genetic alterations. Furthermore, the robustness of our model was enhanced by hyperparameter tuning through a grid search to avoid overfitting and we evaluated its predictive performance using both internal cross-validation and an independent validation cohort ^51^.

Through this improved framework, we successfully prioritized CAD-associated genes, extending beyond previously reported genes such as *LDLR*, *FTO,* and *CYP27A1*. By mapping these genes onto the human protein-protein interaction network, we uncovered 46 tightly clustered topological modules, providing insights into their functional roles in CAD pathogenesis. Beyond lipid metabolism, the analysis revealed modules associated with other relevant biological processes, including platelet function, immune system regulation, blood vessel and heart development, and RNA metabolism. Interestingly, while previous GWASs have highlighted the role of common variants in CAD development through various pathways, our findings suggest that rare variants also contribute to the disease through a wide spectrum of biological processes.

We also utilized our framework to develop an RVS and demonstrated its discriminative capacity between CAD cases and controls in the validation cohort. The distinctive feature of RVS lies in its utilization of rare nonsynonymous variants as input data, setting it apart from conventional PRS that primarily focus on common variants. This approach allows RVS to tap into a different spectrum of genomic information, involving risk factors uncaptured by PRS. The independence of RVS from PRS is further substantiated by the absence of a significant positive correlation between these two scoring systems and the complementary relationships with clinical risk parameters. This lack of correlation suggests that the RVS and PRS are capturing distinct aspects of genetic risk for CAD, each contributing unique information to the overall risk assessment. Importantly, the integration of RVS and PRS resulted in improved predictive performance, demonstrating a synergistic effect that enhanced the ability to accurately assess CAD risk. While methods combining information from one or a few genetic mutations with PRS have been reported ^52^, our study presented a more comprehensive approach to combine rare and common variant information. Furthermore, these findings reinforce the recognition that rare variants, despite their low frequency, contribute significantly to the genetic architecture of CAD and can help explain a portion of its missing heritability that common variants alone cannot account for.

There are several limitations in the study. First, there was a difference in age distribution between cases and controls. This discrepancy arose because we specifically selected early-onset CAD patients for the case group, resulting in a younger average age. As in previous rare variant studies, we prioritized selecting early-onset CAD cases to enrich genetic contributions ^20^. Second, some of the prioritized genes for CAD in this study have unknown functions, especially in cluster 6. However, many loci and genes identified in GWAS on CAD remain functionally uncharacterized, as well ^41,53^. Therefore, future research is necessary to investigate the gene function and biological pathways to CAD development. Third, this study used WGS data from the Japanese population, so it is not certain whether the RVS created in this study can be applied to other populations since a PRS derived from GWAS in one population is reported to be less accurate in other populations ^11,54^. These results need to be validated in other populations and prospective cohorts.

Taken together, our study underscores the important role of rare variants in the genetic landscape of CAD. By leveraging a machine learning-based framework, we have revealed CAD-associated genes and pathways influenced by rare variants. Our results demonstrate the distinct and complementary value of RVS compared to conventional PRS, highlighting the enhanced predictive power achieved through their integration. This comprehensive approach offers new insights into the pathogenesis of CAD, potentially leading to the accurate assessment and management of individual CAD risk.

## Consortia

### The Biobank Japan Project

Koichi Matsuda^1,2^, Takayuki Morisaki^2,3^, Yukinori Okada^4^, Yoichiro Kamatani^5^, Kaori Muto^6^, Akiko Nagai^6^, Yoji Sagiya^2^, Natsuhiko Kumasaka^7^, Yoichi Furukawa^8^, Yuji Yamanashi^3^, Yoshinori Murakami^3^, Yusuke Nakamura^3^, Wataru Obara^9^, Ken Yamaji^10^, Kazuhisa Takahashi^11^, Satoshi Asai^12,13^, Yasuo Takahashi^13^, Shinichi Higashiue^14^, Shuzo Kobayashi^14^, Hiroki Yamaguchi^15^, Yasunobu Nagata^15^, Satoshi Wakita^15^, Chikako Nito^16^, Yu-ki Iwasaki^17^, Shigeo Murayama^18^, Kozo Yoshimori^19^, Yoshio Miki^20^, Daisuke Obata^21^, Masahiko Higashiyama^22^, Akihide Masumoto^23^, Yoshinobu Koga^23^ & Yukihiro Koretsune^24^

^1.^Laboratory of Genome Technology, Human Genome Center, Institute of Medical Science, The University of Tokyo, Tokyo, Japan.

^2^ Laboratory of Clinical Genome Sequencing, Graduate School of Frontier Sciences, The University of Tokyo, Tokyo, Japan.

^3^ The Institute of Medical Science, The University of Tokyo, Tokyo, Japan.

^4^ Department of Genome Informatics, Graduate School of Medicine, The University of Tokyo, Tokyo, Japan.

^5^ Laboratory of Complex Trait Genomics, Graduate School of Frontier Sciences, The University of Tokyo, Tokyo, Japan.

^6^ Department of Public Policy, Institute of Medical Science, The University of Tokyo, Tokyo, Japan.

^7^ Division of Digital Genomics, Institute of Medical Science, The University of Tokyo, Tokyo, Japan.

^8^ Division of Clinical Genome Research, Institute of Medical Science, The University of Tokyo, Tokyo, Japan.

^9^ Department of Urology, Iwate Medical University, Iwate, Japan.

^10^ Department of Internal Medicine and Rheumatology, Juntendo University Graduate School of Medicine, Tokyo, Japan.

^11^ Department of Respiratory Medicine, Juntendo University Graduate School of Medicine, Tokyo, Japan.

^12^ Division of Pharmacology, Department of Biomedical Science, Nihon University School of Medicine, Tokyo, Japan.

^13^ Division of Genomic Epidemiology and Clinical Trials, Clinical Trials Research Center, Nihon University. School of Medicine, Tokyo, Japan.

^14^ Tokushukai Group, Tokyo, Japan.

^15^ Department of Hematology, Nippon Medical School, Tokyo, Japan.

^16^ Laboratory for Clinical Research, Collaborative Research Center, Nippon Medical School, Tokyo, Japan.

^17^ Department of Cardiovascular Medicine, Nippon Medical School, Tokyo, Japan.

^18^ Tokyo Metropolitan Geriatric Hospital and Institute of Gerontology, Tokyo, Japan.

^19^ Fukujuji Hospital, Japan Anti-Tuberculosis Association, Tokyo, Japan.

^20^ The Cancer Institute Hospital of the Japanese Foundation for Cancer Research, Tokyo, Japan.

^21^ Center for Clinical Research and Advanced Medicine, Shiga University of Medical Science, Shiga, Japan.

^22^ Department of General Thoracic Surgery, Osaka International Cancer Institute, Osaka, Japan.

^23^ Iizuka Hospital, Fukuoka, Japan.

^24^ National Hospital Organization Osaka National Hospital, Osaka, Japan.

## Data Availability

All data produced in the present study are available upon reasonable request to the authors.

## Acknowledgements

We thank the staff of BBJ and the Nagahama cohort study for their assistance in collecting samples and clinical information. We thank the participants in the BBJ and Nagahama cohort study for their contribution to the study. H.I. is funded by the Japan Society for the Promotion of Science grant (JP22J00780, JP22K16128). K.I. is supported by the Japan Agency for Medical Research and Development (AMED) under grant numbers JP24bm1423005, JP24km0405209, JP24tm0524004, JP24tm0624002, JP24km0405209 and JP24ek0210164. K.I. and K.O. are supported by the Research Funding for Longevity Sciences from the NCGG (24–15). BBJ is supported by the Tailor-Made Medical Treatment Program of the Ministry of Education, Culture, Sports, Science, and Technology (MEXT) and AMED under grant numbers JP17km0305002 and JP17km0305001, JP.24tm0624002. The Nagahama study was supported by a JSPS Grant-in-Aid for Scientific Research (C), KAKENHI grant numbers JP17K07255 and JP17KT0125, and the Practical Research Project for Rare/Intractable Diseases from AMED under grant numbers JP16ek0109070, JP18kk0205008, JP18kk0205001, JP19ek0109283, and JP19ek0109348.

## Author contributions

H.I. and K.I. conceived and designed the study. C.K., J.S., K.H., and F.M. collected, managed and genotyped the Nagahama cohort. K.M., C.T. and Y.K. collected and managed the BBJ samples. H.I. and K.I. analyzed WGS data, developed the machine-learning model and performed the statistical analyses. S. K estimated the effect size to calculate PRS. S.Z. developed the PPI network module and analyzed it. H.Y., R.K., H.M., K.M., N.E., K.O., Y.O., C.T., and Y.K. contributed to data processing, analysis and interpretation. K.I., M.S. and I.K. supervised the study. H.I. and K.I. wrote the manuscript, and many authors have provided valuable insights and edits.

## Supplemental information

**Document S1.** Table S1-S3, S5, S7, S9-13, Figure S1-8

**Table S4.** Summary statistics of aggregated rare variant association analysis using SAIGE-GENE+, related to Figure S4.

**Table S6.** Summary of 59 HEAL_CAD_ genes with gene-based annotation, related to Figure 2.

**Table S8.** The 46 Protein Interaction Modules Identified in CAD, Related to Figure 2.

## STAR Methods

### Code availability

The code of the modified HEAL framework is available on https://github.com/pirocv/HEAL.

### Study cohort

Two previously described cohorts were used in the current study. BioBank Japan (BBJ) is a hospital-based Japanese biobank project including clinical and genetic data from a variety of patients ^55,56^. Participants were recruited from 12 hospitals throughout Japan. The Nagahama Prospective Genome Cohort (Nagahama study) is the genome cohort conducted in Shiga, Japan. Participants aged 30–74 years were recruited from the general population in Nagahama city from 2007 to 2010 ^57^.

### Whole genome sequencing and quality control

We sequenced 1,765 CAD patients and 3,148 controls from the cohort. Whole genome sequence (WGS) was performed on Illumina’s HiSeqX aiming at 15x depth, using 150-base pair-end reads. We also sequenced an additional 200 CAD cases and 836 controls aiming at 30x depth using 150-base paired-end reads. In order to enrich for a genetic contribution to disease ^20^, we prioritized patients with early-onset MI, one of the most severe forms of CAD, within the BBJ cohort for WGS (age of MI onset in 15x and 30x WGS cohort: 47.4 ± 4.1 years and 36.0 ± 3.9 years, respectively). Sequenced reads were aligned to the hs37d5 reference genome using BWA software ^58^. The genotypes of the samples were called using the HaplotypeCaller implemented in GATK v3.8. Per-sample Genomic Variant Call Format (gVCF) genotype data were merged and jointly called using GenotypeGVCFs. We defined exclusion filters for genotypes as follows. (1) For 15x depth data, filtered depth (DP) < 2, quality of the assigned genotype (genotype quality; GQ) < 20. (2) For 30x depth data, DP < 5, GQ < 20, DP > 60 and GQ < 95. We set these genotypes as missing and excluded variants with call rates < 90% before variant quality score recalibration. For sample quality control, the following samples were excluded: (1) age < 20 years old, (2) excess missing genotypes (> 10%), (3) samples whose genetically inferred sex did not match the self-reported sex, (4) closely related samples estimated by identity-by-descent and identity-by-state analysis (Pi-hat > 0.1875) and (5) excess heterozygosity. We also excluded non-Japanese participants estimated from Principal component analysis (PCA) calculated using PLINK 2.0 ^59^. The total number of genomes that failed data quality control is summarized in **Table S13**. After the sample quality control, we retained 1,752 CAD case samples and 3019 non-CAD control samples for 15x depth data and 200 case samples and 824 control samples for 30x depth. Then, the variant quality control was performed excluding (1) high missingness (5% for 15x depth and 1% for 30x depth), (2) Hardy-Weinberg equilibrium (P < 1 *10^-6^), (3) variants in the low complexity region. WGS data with 15x depth data was used as a discovery cohort and the 30x depth data was used as the validation cohort in the machine learning-based analysis.

### Single variant association analysis

The single variant association test was performed by logistic regression implemented in PLINK 2.0 ^60^ with adjustment for age, sex, and the first 10 principal components of ancestry. Principal components of ancestry were calculated using PLINK 2.0 ^59^. The inclusion of principal components as covariates in the logistic regression analysis increases the power to detect true genetic associations and minimizes confounding by population stratification ^61^. Variants with a missing rate of less than 0.01 were included in the analysis. Genomic inflation factor (λ*_GC_*) was calculated using variants with MAF ≥ 0.001. Single variant association analysis was also performed using SAIGE ^62^ with adjustment for age, sex, and the first 10 principal components of ancestry. SAIGE is widely used in GWASs for binary traits to account for population structure and relatedness while correcting for the type I error rates ^62^. The genome-wide significance threshold was set at P = 5 * 10^-8^. To define a locus, we added 500 kb to both sides of each genome-wide significant SNP and merged overlapping regions. To determine whether each locus was novel, a literature search was conducted to ascertain if any of the regions contained SNPs had been previously reported as significant for CAD.

### Aggregated rare variant association analysis

We also performed gene-based association analysis using SAIGE-GENE+ software, which accounts for the relatedness among the study samples ^63,64^. We first calculated sparse GRM using the WGS data and fit the null model in the SAIGE-GENE+ algorithm step1. For the gene-based association analysis, we extracted rare (MAF < 0.001) nonsynonymous variants including (nonsynonymous single nucleotide variations (SNV), nonframeshift insertion, nonframeshift deletion, frameshift insertion, frameshift deletion, stopgain, stoploss, and splice site variants). Splice-site variants, pLOF variants and damaging missense variants defined by a REVEL score > 0.5 ^65^ were included in the analysis. SKAT-O test implemented in SAIGE-GENE+ software was performed with adjustment for age, sex and first 10 principal components of ancestry. Gene-wide significance threshold and suggestive threshold were set at P = 2.5 * 10^-6^ and P = 5 * 10^-4^, respectively. Statistical inflation was estimated by Q-Q plot.

### Machine learning-based analysis (modified HEAL)

We employed a recently developed machine learning-based rare variant analysis method called HEAL (hierarchical estimate from agnostic learning). A detailed HEAL method is described in the original paper ^19^. In this framework (**Figure S8**), we first annotated each variant using ANNOVAR software ^66^ and extracted rare nonsynonymous variants (nonsynonymous SNV, nonframeshift insertion, nonframeshift deletion, frameshift insertion, frameshift deletion, stopgain, stoploss, and splice site variants) that were not present in the East-Asian populations analyzed in the 1000 Genomes Project ^67^. Variants with high frequency in the WGS data and gnomAD East Asian database ^68^ (MAF > 0.1) were also filtered. To estimate the mutation burden for each gene based on the rare variants, we used the REVEL score (ranges from 0 to 1 with a higher score indicating a damaging variant), which was internally computed by ANNOVAR software. The deleteriousness score of the putative loss of function (pLOF) variants, such as stopgain and splice site variants, was set as 1. Next, we calculated the cumulative effects of rare nonsynonymous variants for each gene as

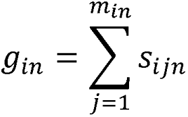

where *g_in_* is the mutation burden of the gene *i* of *n*th sample, *m_in_* is the number of rare nonsynonymous variants, *s_ijn_* is the deleteriousness score for variant *j* of gene *i*. Using the above formula, we obtained a matrix of estimated mutation burden for each gene per sample (*x_n_* = (*g*_l*n*_,*g*_2*n*_,…,*g_mn_*), where m is the number of the total genes). The mutation burden was standardized (Z-score normalization). We trained a regularized logistic regression model for a genome-based CAD prediction model. The input of the model is the calculated mutation burden and the output is the probability of CAD as shown in the following equation.

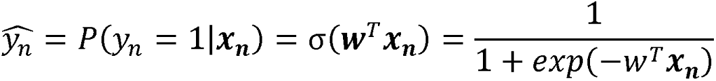

where *y_n_* is the label for CAD case (1) or control (0), 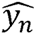 is the probability of being CAD positive given the mutation burden ***x_n_*** for the *n*th sample, σ is the sigmoid function and ***w*** is the weight vector. To identify the optimal coefficient vector ***w*** that achieve the maximum consistency between the model probabilities (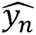) and the observations for the cohort (*y_n_*), we solved the following optimization problem.

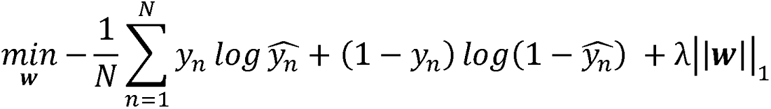

In this regularized logistic regression, regularization strength is determined by parameter λ, and it is a hyperparameter of the machine learning model, which was determined by the cross-validation method (**Figure S5**). By training the model to predict disease status, it outputs the minimal set of most distinguishing features (genes) for CAD. The trained model can be used to estimate the rare variant-based disease risk score (RVS) from the genomic data. We have named this the modified HEAL because our approach differs from the original method in that we included not only missense variants but also pLOF variants. We determined the hyperparameters using grid search and estimated the performance in the independent cohort to avoid bias and overestimation of the model’s performance, while the performance was estimated using internal cross-validation in the original method.

### Interpretation of genes identified by modified HEAL

To investigate the functions of the 59 identified genes, we first annotated each one using various databases and then conducted clustering analysis to categorize the groups of genes to obtain the eight functional groups. Annotations included checking the constraint score (pLI) from the gnomAD database ^68^, identifying whether the genes were reported in previous GWAS on CAD and its risk factors (lipids, diabetes, obesity, blood pressure, coagulation function, and smoking-related phenotypes) using the GWAS Catalog, and checking for the overlap with target genes of enhancers that were significant in previous GWAS on CAD and its risk factors (same as above) using the GeneHancer database, which includes genome-wide enhancers and their target genes ^21^. Further analysis involved examining the International Mouse Phenotyping Consortium (IMPC) database to determine if the corresponding genes in knock-out mice are significantly related to phenotypes such as blood pressure, blood glucose and lipid traits. Enrichment analysis for Gene Ontology and Human Phenotype Ontology was performed using g:Profiler ^69^ to gain insights into the biological processes and human phenotypic abnormalities associated with these genes ^22^. We considered statistical significance for the enrichment analysis with a false discovery rate under 0.1.

To analyze the functional modules in CAD, we downloaded the human protein-protein interactions (PPIs) from STRING v12.0, comprising 19,622 proteins and 6,857,702 interactions. High-confidence PPIs (combined score >700) were extracted for downstream analysis, including 16,185 proteins and 236,000 interactions. To remove bias from hub proteins, we applied the random walk with restart (RWR) algorithm with a restart probability of 0.5. This produced a smoothed network after retaining the top 5% predicted edges (n = 6,243,766). We employed the Louvain method ^70^ to decompose the network into different modules. Following algorithm convergence, we obtained 1,261 modules with an average size of 13 nodes. Among the 1,261 PPI modules, 46 encompassed at least one gene identified by the machine learning analysis. We used g:profiler to determine functional enrichment for each module. Cytoscape software ^71^ was used to visualize the PPI modules.

### Genetic risk scores

By optimizing the machine learning-based model, the modified HEAL framework can also make a prediction of disease based on the input genome. We call it rare variant-based genetic risk score (RVS) because it only leverages information on rare variants. Using the trained model, we estimated the RVS prediction performance in the validation cohort. We also analyzed the association between RVS and clinical parameters such as vital signs and blood test data in the BBJ data using Pearson’s correlation. To investigate the prognostic impact of RVS, we divided the patients into those in the top 5% and those below, then compared their outcome using Kaplan-Meier analysis and a log-rank test. To compare the properties between RVS and the common variant-based polygenic risk score (PRS), GWAS of CAD in BBJ (case 25,668 vs control 141,667) was performed. The individuals included in the GWAS were genotyped using the HumanOmniExpressExome v.1.0/v.1.2 platform (Illumina) or in combination with HumanOmniExpress v.1.0 and Human Exome BeadChip v.1.0/v.1.1 (Illumina). For genotype quality control, variants with (1) SNP call rate < 99%, (2) Hardy–Weinberg equilibrium (P < 1 *10^-6^) and (3) heterozygous counts <5 were excluded. We performed pre-phasing using Eagle software. Phased haplotypes were imputed to the in-house reference panel from BBJ ^11^ by minimac3 ^72^. Variants with low imputation quality (R^2^<0.3) were excluded. GWAS was performed by logistic regression implemented in PLINK 2.0 ^60^ with adjustment for age, age^2^, sex and first 10 principal components of ancestry. Then PRS of *i*th sample was calculated as follows

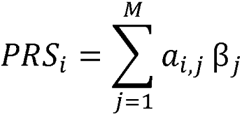

where M is the number of variants in GWAS, *a_i,j_* is the number of effect allele of *j*th variant in *i*th sample, and β*_j_* is the effect size of *j*th variant estimated by GWAS. The number of variants included in the PRS calculation was determined by the pruning and thresholding method ^13^. The relationship between RVS and PRS was examined by Pearson’s correlation coefficient, both in cases only and across the validation cohort. We then integrated both RVS and PRS by normalizing (mean 0, standard deviation 1) and adding them together to obtain combined risk score (CRS). The predictive performance of each genetic score was estimated on the validation cohort, which was not used in the derivation of the RVS, PRS, or CRS. We used receiver operating characteristics (ROC) to evaluate the predictive performance. To examine whether CRS improves predictive performance compared to conventional PRS, we compared AUROC of PRS and CRS by DeLong’s test. We also calculated the area under precision-recall curve (AUPRC) and Nagelkerke’s pseudo R^2^ metrics. The P values were derived using a 20000 times bootstrap replication method. In all statistical analyses, R software was used and a two-sided P < 0.05 was considered statistically significant.

