## Supplementary document S1 for "Machine Learning Reveals the Contribution of Rare Genetic Variants and Enhances Risk Prediction for Coronary Artery Disease in the Japanese Population"

**Supplementary Tables**

**Table S1. The number of individuals before and after quality control in this study.**

| Data | WGS target depth | Disease status | Data source | N (After QC) | N (pre-QC) |
| --- | --- | --- | --- | --- | --- |
| Discovery cohort | 15x | Case | BBJ | **1,752** | 1765 |
|  |  | Control | BBJ + Nagahama | **3,019** | 3148 |
| Validation cohort | 30x | Case | BBJ | **200** | 200 |
|  |  | Control | BBJ | **824** | 836 |

The number of Individuals and their sources included in the study before and after sample quality control are listed. BBJ, BioBank Japan; QC, quality control.

**Table S2. Breakdown of variants in the datasets by the function of the variants**

| Data | Total number of variants | SNV | Insertions | Deletions |
| --- | --- | --- | --- | --- |
| Discovery cohort | 51,717,580 | 48,489,965 | 2,284,448 | 943,167 |
| Validation cohort | 25,531,471 | 23,950,464 | 490,513 | 619,485 |

The number of variants included in the discovery and validation cohort data. SNV, single nucleotide variant.

**Table S3. GWAS significant loci in SAIGE analysis (related to Figure S3).**

| rs ID | CHR | POS | Allele1 | Allele2 | AF_Allele2 | BETA | SE | P value | Known region |
| --- | --- | --- | --- | --- | --- | --- | --- | --- | --- |
| rs7977233 | 12 | 109753314 | T | C | 0.164536 | 0.407718 | 0.07226 | **1.47E-08** | YES |
| rs3782886 | 12 | 112110489 | T | C | 0.28799 | 0.334 | 0.059977 | **2.40E-08** | YES |

Summary statistics of the two loci that reached genome-wide significance threshold in the single variant association analysis using SAIGE software. CHR, chromosome; POS, position; AF, allele frequency; SE, standard deviation

**Table S5. A minimal set of 59 HEAL_CAD_ genes that are associated with CAD (related to Figure 2 and Figure S6).**

| *LDLR* | *SHROOM3* |
| --- | --- |
| *CCT6B* | *KLF9* |
| *VRK2* | *MTO1* |
| *PIWIL2* | *SIVA1* |
| *WBP2* | *PRSS3* |
| *WNT10A* | *GALK1* |
| *TSSK1B* | *CLDN14* |
| *RNF216* | *TSPAN14* |
| *SLC35G6* | *COMMD10* |
| *FTO* | *CPOX* |
| *EGFLAM* | *RGSL1* |
| *GIT1* | *SPOPL* |
| *KDR* | *DIS3L2* |
| *OR6X1* | *CLN6* |
| *ST3GAL5* | *VPS4B* |
| *CYP27A1* | *REM1* |
| *FAM193B* | *SEC14L1* |
| *ZBTB18* | *DOCK2* |
| *PLA2G16* | *RBM12* |
| *OPTN* | *PAM16* |
| *ZNF25* | *LUC7L3* |
| *TFF2* | *SMYD5* |
| *RPUSD2* | *SLC27A5* |
| *CIDEC* | *SLC7A11* |
| *SLC12A4* | *TFPI2* |
| *BOLL* | *PIH1D2* |
| *FREM3* | *DCP2* |
| *LRRC31* | *CEP19* |
| *DERL1* | *MYO6* |
| *NTRK3* |  |

**Table S7. Gene set enrichment analysis of HEAL_CAD_ genes (related to Figure 2).**

| Type | Name | # Genes in Gene Set (K) | Description | # Genes in Overlap (k) | k/K | p-value | FDR q-value |
| --- | --- | --- | --- | --- | --- | --- | --- |
| GOMF | PROTEIN_CONTAINING_COMPLEX_BINDING | 1281 | Binding to a macromolecular complex. [GOC:jl] | 11 | 0.0086 | 1.33E-06 | 2.30E-02 |
| HP | PREMATURE_CORONARY_ARTERY_ATHEROSCLEROSIS | 18 | Premature coronary artery atherosclerosis | 3 | 0.1667 | 2.04E-06 | 2.30E-02 |
| GOBP | CELLULAR_MACROMOLECULE_CATABOLIC_PROCESS | 1055 | The chemical reactions and pathways resulting in the breakdown of a macromolecule, any large molecule including proteins, nucleic acids and carbohydrates, as carried out by individual cells. [GOC:jl] | 9 | 0.0085 | 1.43E-05 | 7.08E-02 |
| HP | ABNORMAL_CIRCULATING_CHOLESTEROL_CONCENTRATION | 112 | Abnormal circulating cholesterol concentration | 4 | 0.0357 | 1.86E-05 | 7.08E-02 |
| GOBP | NEGATIVE_REGULATION_OF_RECEPTOR_RECYCLING | 5 | Any process that stops, prevents, or reduces the rate of receptor recycling. [GOC:add] | 2 | 0.4 | 1.89E-05 | 7.08E-02 |
| GOBP | REGULATION_OF_HISTONE_H3_K14_ACETYLATION | 5 | Any process that modulates the frequency, rate or extent of the addition of an acetyl group to histone H3 at position 14 of the histone. [GOC:mah] | 2 | 0.4 | 1.89E-05 | 7.08E-02 |
| GOBP | MACROMOLECULE_CATABOLIC_PROCESS | 1408 | The chemical reactions and pathways resulting in the breakdown of a macromolecule, any molecule of high relative molecular mass, the structure of which essentially comprises the multiple repetition of units derived, actually or conceptually, from molecules of low relative molecular mass. [GOC:mah] | 10 | 0.0071 | 2.21E-05 | 7.10E-02 |
| GOCC | SORTING_ENDOSOME | 6 | A multivesicular body surrounded by and connected with multiple tubular compartments with associated vesicles. [NIF_Subcellular:sao1028571114] | 2 | 0.3333 | 2.83E-05 | 7.32E-02 |
| HP | INCREASED_BLOOD_PRESSURE | 429 | Increased blood pressure | 6 | 0.014 | 2.93E-05 | 7.32E-02 |
| HP | ABNORMAL_CIRCULATING_LIPID_CONCENTRATION | 282 | Abnormal circulating lipid concentration | 5 | 0.0177 | 4.64E-05 | 9.59E-02 |

GOMF, gene ontology molecular functions; GOBP, gene ontology biological process; GOCC, gene ontology Cellular Components.

**Table S9. Correlation between RVS and clinical measurements (related to Figure 3).**

| Clinical measurements | Pearson's r | Lower 95% C.I. | Upper 95% C.I. | p value |
| --- | --- | --- | --- | --- |
| LDLC | 0.211 | 0.111 | 0.307 | 4.54E-05 |
| TBil | 0.134 | 0.070 | 0.198 | 4.98E-05 |
| ALT | 0.126 | 0.064 | 0.187 | 7.16E-05 |
| PTINR | 0.097 | 0.009 | 0.183 | 3.05E-02 |
| TC | 0.093 | 0.025 | 0.160 | 7.21E-03 |
| K | 0.068 | 0.004 | 0.131 | 3.75E-02 |
| Hb | 0.062 | 0.000 | 0.124 | 5.18E-02 |
| UA | 0.061 | -0.013 | 0.134 | 1.06E-01 |
| APTT | 0.059 | -0.034 | 0.151 | 2.11E-01 |
| Alb | 0.059 | -0.009 | 0.126 | 8.77E-02 |
| RBC | 0.058 | -0.004 | 0.120 | 6.77E-02 |
| AST | 0.058 | -0.005 | 0.120 | 6.94E-02 |
| WBC | 0.048 | -0.015 | 0.110 | 1.36E-01 |
| CK | 0.048 | -0.030 | 0.125 | 2.31E-01 |
| TP | 0.043 | -0.022 | 0.108 | 1.93E-01 |
| Cre | 0.035 | -0.027 | 0.097 | 2.71E-01 |
| DBP | 0.035 | -0.032 | 0.101 | 3.08E-01 |
| SBP | 0.026 | -0.041 | 0.092 | 4.51E-01 |
| BUN | 0.025 | -0.038 | 0.088 | 4.33E-01 |
| TG | 0.023 | -0.048 | 0.094 | 5.29E-01 |
| CRP | 0.019 | -0.062 | 0.099 | 6.53E-01 |
| PLT | 0.016 | -0.048 | 0.079 | 6.29E-01 |
| P | 0.011 | -0.099 | 0.121 | 8.42E-01 |
| cGTP | -0.002 | -0.069 | 0.064 | 9.45E-01 |
| BS | -0.006 | -0.078 | 0.066 | 8.64E-01 |
| LDH | -0.009 | -0.074 | 0.056 | 7.86E-01 |
| Na | -0.011 | -0.076 | 0.053 | 7.31E-01 |
| Ca | -0.030 | -0.111 | 0.051 | 4.65E-01 |
| HbA1c | -0.032 | -0.134 | 0.071 | 5.42E-01 |
| ALP | -0.038 | -0.107 | 0.031 | 2.77E-01 |
| Cl | -0.043 | -0.108 | 0.022 | 1.96E-01 |
| HDLC | -0.069 | -0.157 | 0.020 | 1.28E-01 |

Correlation between RVS and continuous clinical indices. C.I., confidence interval; LDLC, low-density lipoprotein cholesterol; Tbil, total bilirubin; ALT, alanine aminotransferase; PTINR, prothrombin time international normalized ratio; TC, total cholesterol; K, potassium; Hb, hemoglobin; UA, uric acid; APTT, activated partial thromboplastin time; Alb, albumin; RBC, red blood cell; AST, aspartate aminotransferase; WBC, white blood cell; CK, creatine kinase; TP, total protein; Cre, creatinine; DBP, diastolic blood pressure; SBP, systolic blood pressure; BUN, blood urea nitrogen; TG, triglycerides; CRP, C-reactive protein; PLT, platelet; P, Phosphorus; γGTP, gamma-glutamyl transpeptidase; BS, blood sugar; LDH, Lactate dehydrogenase.

**Table S10. Correlation between PRS and clinical measurements (related to Figure S7).**

| Clinical measurements | Pearson's r | Lower 95% C.I. | Upper 95% C.I. | p value |
| --- | --- | --- | --- | --- |
| UA | 0.147 | 0.075 | 0.218 | 8.18E-05 |
| APTT | 0.127 | 0.035 | 0.217 | 7.19E-03 |
| TG | 0.117 | 0.046 | 0.186 | 1.22E-03 |
| LDH | 0.083 | 0.017 | 0.147 | 1.31E-02 |
| BS | 0.081 | 0.009 | 0.152 | 2.76E-02 |
| TP | 0.070 | 0.005 | 0.135 | 3.37E-02 |
| CK | 0.059 | -0.019 | 0.136 | 1.37E-01 |
| Ca | 0.059 | -0.023 | 0.139 | 1.58E-01 |
| Hb | 0.055 | -0.008 | 0.117 | 8.65E-02 |
| RBC | 0.052 | -0.011 | 0.114 | 1.03E-01 |
| Alb | 0.051 | -0.017 | 0.118 | 1.42E-01 |
| cGTP | 0.049 | -0.017 | 0.115 | 1.47E-01 |
| SBP | 0.044 | -0.023 | 0.110 | 2.01E-01 |
| WBC | 0.038 | -0.025 | 0.100 | 2.37E-01 |
| AST | 0.037 | -0.025 | 0.099 | 2.46E-01 |
| HbA1c | 0.030 | -0.073 | 0.132 | 5.74E-01 |
| Cre | 0.023 | -0.039 | 0.086 | 4.64E-01 |
| DBP | 0.023 | -0.044 | 0.090 | 4.95E-01 |
| ALT | 0.020 | -0.043 | 0.082 | 5.32E-01 |
| K | 0.015 | -0.049 | 0.079 | 6.35E-01 |
| TBil | 0.014 | -0.052 | 0.079 | 6.83E-01 |
| TC | 0.008 | -0.059 | 0.076 | 8.07E-01 |
| BUN | 0.000 | -0.063 | 0.064 | 9.91E-01 |
| ALP | -0.002 | -0.071 | 0.067 | 9.51E-01 |
| PTINR | -0.008 | -0.095 | 0.080 | 8.66E-01 |
| Na | -0.018 | -0.082 | 0.047 | 5.91E-01 |
| PLT | -0.034 | -0.097 | 0.029 | 2.92E-01 |
| Cl | -0.035 | -0.100 | 0.030 | 2.92E-01 |
| CRP | -0.044 | -0.124 | 0.036 | 2.82E-01 |
| LDLC | -0.061 | -0.162 | 0.042 | 2.45E-01 |
| P | -0.064 | -0.173 | 0.046 | 2.53E-01 |
| HDLC | -0.100 | -0.188 | -0.011 | 2.73E-02 |

Correlation between PRS and continuous clinical indices. C.I., confidence interval; LDLC, low density lipoprotein cholesterol; Tbil, total bilirubin; ALT, alanine aminotransferase; PTINR, prothrombin time international normalized ratio; TC, total cholesterol; K, potassium; Hb, hemoglobin; UA, uric acid; APTT, activated partial thromboplastin time; Alb, albumin; RBC, red blood cell; AST, aspartate aminotransferase; WBC, white blood cell; CK, creatine kinase; TP, total protein; Cre, creatinine; DBP, diastolic blood pressure; SBP, systolic blood pressure; BUN, blood urea nitrogen; TG, triglycerides; CRP, C-reactive protein; PLT, platelet; P, Phosphorus; γGTP, gamma-glutamyl transpeptidase; BS, blood sugar; LDH, Lactate dehydrogenase.

**Table S11. Correlation between CRS and clinical measurements (related to Figure 4).**

| Clinical measurements | Pearson's r | Lower 95% C.I. | Upper 95% C.I. | p value |
| --- | --- | --- | --- | --- |
| UA | 0.148 | 0.076 | 0.220 | 7.18E-05 |
| APTT | 0.134 | 0.041 | 0.223 | 4.62E-03 |
| LDLC | 0.129 | 0.027 | 0.229 | 1.31E-02 |
| ALT | 0.103 | 0.041 | 0.165 | 1.16E-03 |
| TBil | 0.102 | 0.038 | 0.166 | 2.02E-03 |
| TG | 0.099 | 0.028 | 0.168 | 6.39E-03 |
| Hb | 0.083 | 0.021 | 0.144 | 9.22E-03 |
| TP | 0.080 | 0.015 | 0.144 | 1.54E-02 |
| RBC | 0.078 | 0.016 | 0.140 | 1.39E-02 |
| Alb | 0.078 | 0.010 | 0.144 | 2.42E-02 |
| CK | 0.077 | -0.001 | 0.153 | 5.36E-02 |
| TC | 0.073 | 0.005 | 0.140 | 3.53E-02 |
| AST | 0.067 | 0.005 | 0.129 | 3.43E-02 |
| PTINR | 0.064 | -0.024 | 0.151 | 1.55E-01 |
| WBC | 0.061 | -0.002 | 0.123 | 5.74E-02 |
| K | 0.059 | -0.004 | 0.123 | 6.83E-02 |
| BS | 0.054 | -0.018 | 0.126 | 1.40E-01 |
| LDH | 0.053 | -0.013 | 0.118 | 1.14E-01 |
| SBP | 0.050 | -0.017 | 0.116 | 1.46E-01 |
| Cre | 0.042 | -0.021 | 0.104 | 1.93E-01 |
| DBP | 0.041 | -0.025 | 0.108 | 2.25E-01 |
| cGTP | 0.033 | -0.033 | 0.099 | 3.27E-01 |
| BUN | 0.018 | -0.045 | 0.081 | 5.74E-01 |
| Ca | 0.018 | -0.064 | 0.099 | 6.71E-01 |
| HbA1c | -0.003 | -0.105 | 0.100 | 9.59E-01 |
| PLT | -0.013 | -0.077 | 0.050 | 6.79E-01 |
| CRP | -0.019 | -0.099 | 0.062 | 6.46E-01 |
| Na | -0.021 | -0.085 | 0.044 | 5.32E-01 |
| ALP | -0.029 | -0.097 | 0.040 | 4.12E-01 |
| P | -0.035 | -0.145 | 0.075 | 5.33E-01 |
| Cl | -0.055 | -0.120 | 0.010 | 9.49E-02 |
| HDLC | -0.124 | -0.210 | -0.035 | 6.42E-03 |

Correlation between CRS and continuous clinical indices. C.I., confidence interval; LDLC, low density lipoprotein cholesterol; Tbil, total bilirubin; ALT, alanine aminotransferase; PTINR, prothrombin time international normalized ratio; TC, total cholesterol; K, potassium; Hb, hemoglobin; UA, uric acid; APTT, activated partial thromboplastin time; Alb, albumin; RBC, red blood cell; AST, aspartate aminotransferase; WBC, white blood cell; CK, creatine kinase; TP, total protein; Cre, creatinine; DBP, diastolic blood pressure; SBP, systolic blood pressure; BUN, blood urea nitrogen; TG, triglycerides; CRP, C-reactive protein; PLT, platelet; P, Phosphorus; γGTP, gamma-glutamyl transpeptidase; BS, blood sugar; LDH, Lactate dehydrogenase.

**Table S12. Comparison of the predictive performance of RVS, PRS and CRS for CAD in terms of AUROC (related to Figure 5).**

|  | AUROC | | | | AUPRC | | | | Nagelkerke's pseudo R2 | | | |
| --- | --- | --- | --- | --- | --- | --- | --- | --- | --- | --- | --- | --- |
|  | score | C.I.  lower 95% | C.I.  upper 95% | p value  (CRS vs PRS) | score | C.I.  lower 95% | C.I.  upper 95% | p value  (CRS vs PRS) | score | C.I.  lower 95% | C.I.  upper 95% | p value  (CRS vs PRS) |
| RVS | 0.576 | 0.531 | 0.622 | NA | 0.306 | 0.242 | 0.366 | NA | 0.051 | 0.013 | 0.086 | NA |
| PRS | 0.609 | 0.565 | 0.653 | NA | 0.285 | 0.226 | 0.339 | NA | 0.040 | 0.008 | 0.070 | NA |
| CRS | 0.665 | 0.622 | 0.707 | 8.00E-04 | 0.349 | 0.282 | 0.412 | 0.0154 | 0.093 | 0.047 | 0.137 | 0.0018 |

The comparison of the performance of three genetic risk scores for CAD prediction. The confidence intervals (C.I.) were estimated from a 20,000 times bootstrap replication method. AUROC, area under the receiver operating characteristic; AUPRC, area under the precision-recall curve; RVS, rare variant risk score; PRS, polygenic risk score; CRS, combined risk score.

**Table S13. Total number of genomes that failed data quality control (related to Figure 1).**

| Failure reason | Count (with overlap) | |
| --- | --- | --- |
|  | Discovery cohort | Validation cohort |
| Age < 20 years | 5 | 4 |
| Sex ambiguity | 29 | 4 |
| Genetic PCA outlier | 73 | 3 |
| Inbreeding coefficient | 37 | 0 |
| Excess missingness | 3 | 0 |
| Excess heterozygosity | 15 | 3 |
| Total | 142 | 12 |

The term 'with overlap' indicates that individuals may be counted in multiple categories. For example, a genome with both sex ambiguity and excess missingness would be counted in both categories, contributing to the total count in each.

**Supplementary Figures**

**Figure S1 Data and quality control flowchart in the current study**

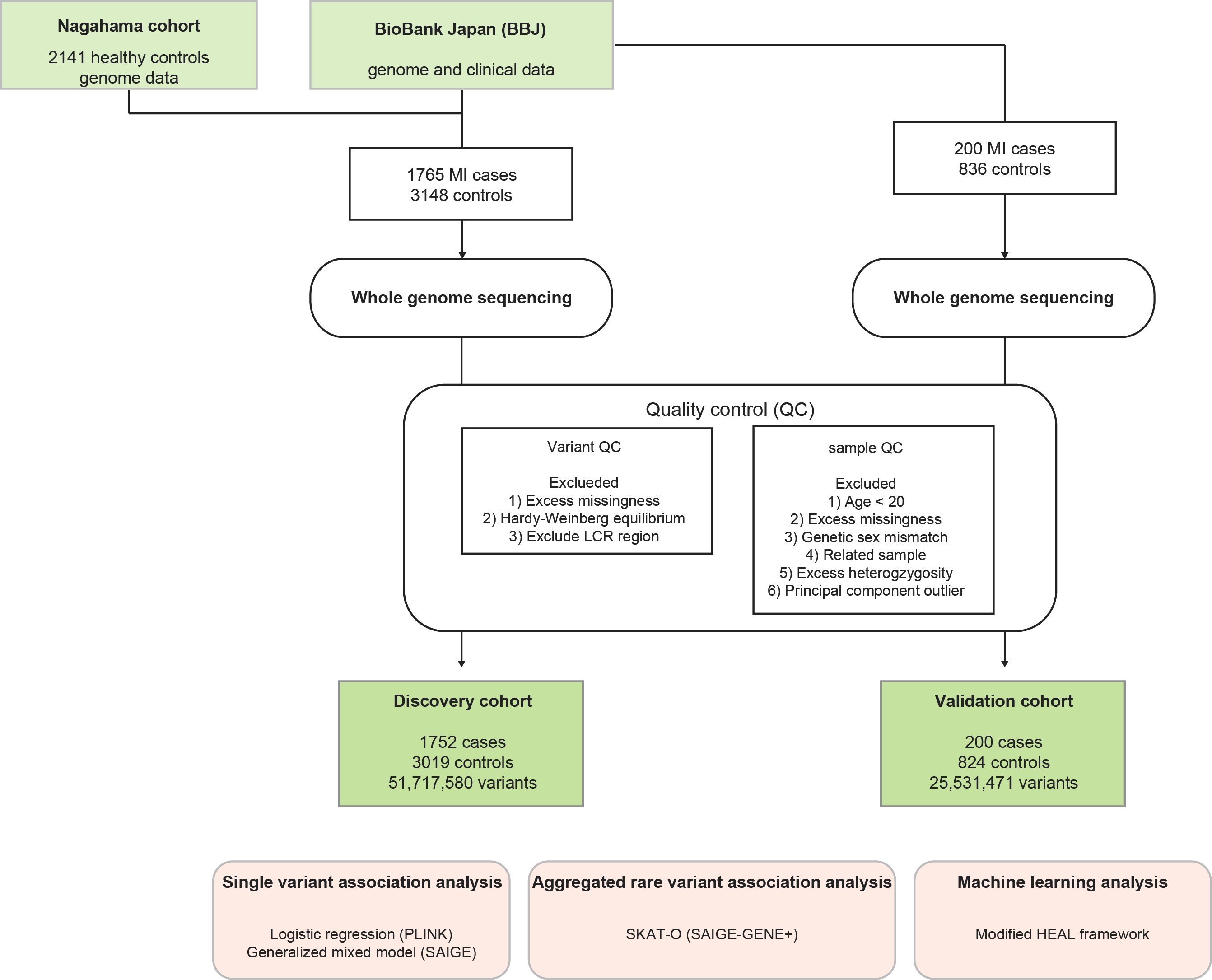
Summary of cohort data and quality control process. We sequenced the whole genome of 1,765 Japanese early-onset MI patients and 3,148 controls for the discovery cohort and 200 MI cases and 836 controls for the validation cohort. After stringent variant and sample-level quality control, single variant association analyses, a gene-based association analysis, and analysis using the modified HEAL framework were performed. MI, myocardial infarction

**Figure S2. Single variant association analysis for CAD by PLINK**

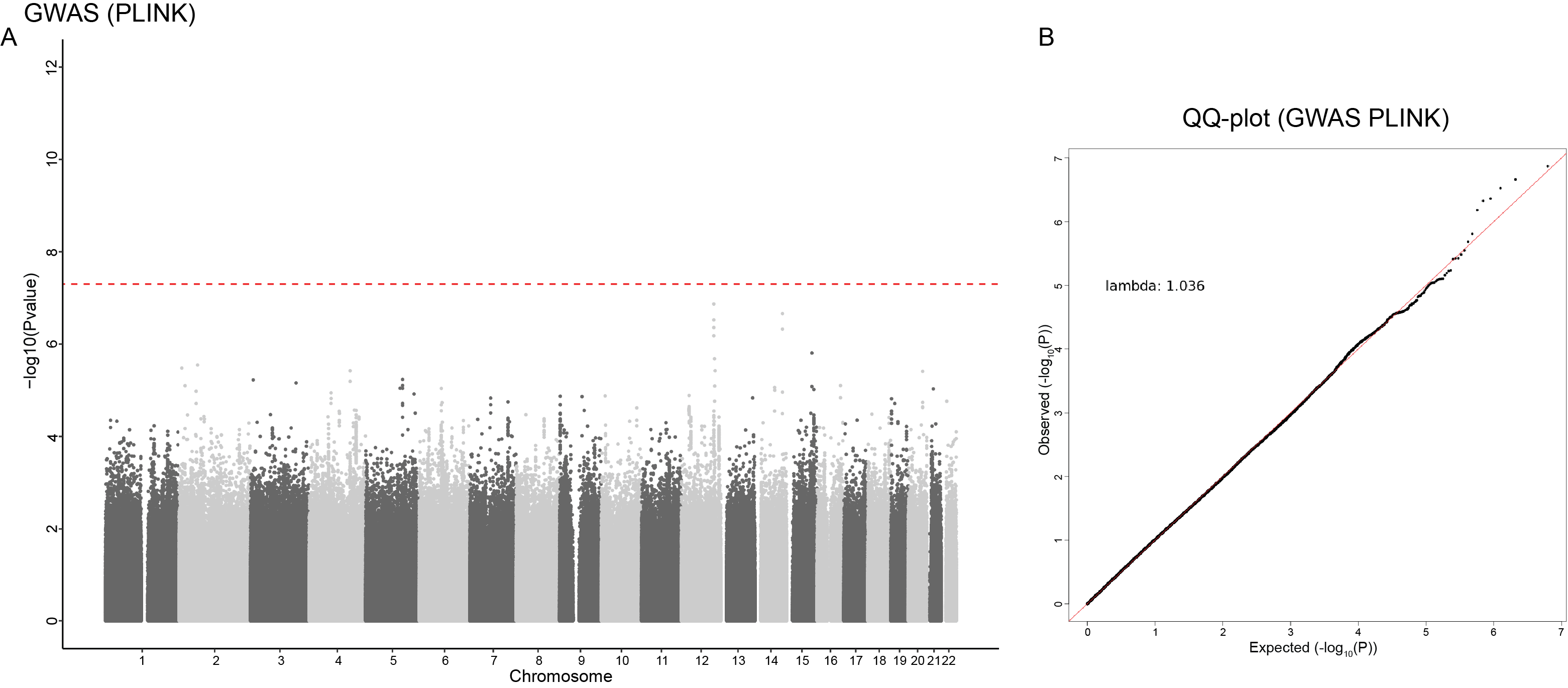

Manhattan plot (A) and QQ plot (B) of the GWAS results for CAD in the discovery cohort of 1752 cases and 3019 controls using PLINK software. The red dashed line represents a genome-wide significance threshold of 5 x 10^-8^.

**Figure S3. Single variant association analysis for CAD by SAIGE.**

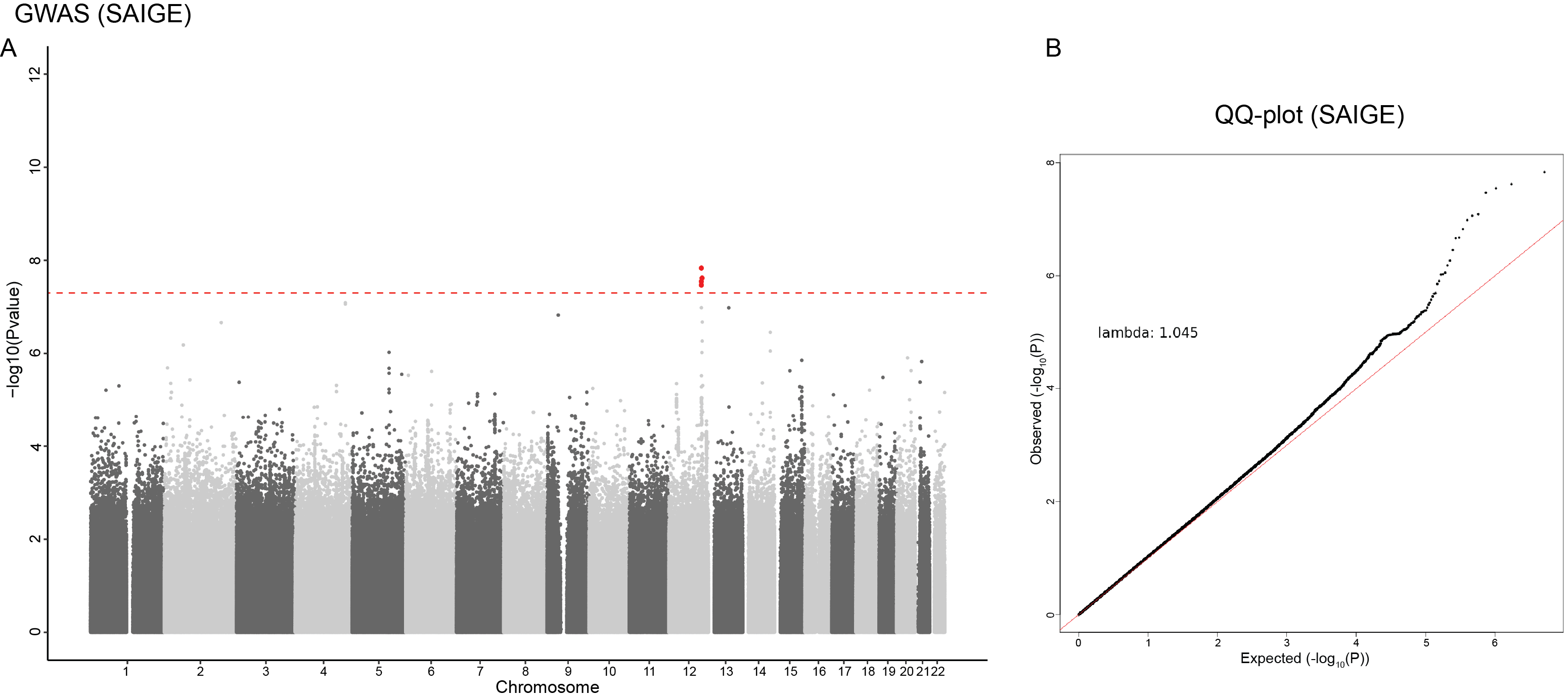

Manhattan plot (A) and QQ plot (B) of the GWAS results for CAD in the discovery cohort of 1752 cases and 3019 controls using SAIGE software. Two loci on chromosome 12 reached a genome-wide significant threshold of 5 x 10^-8^. The red dashed line represents the genome-wide significance threshold.

**Figure S4. Aggregated rare variant association analysis for CAD by SAIGE-GENE+.**

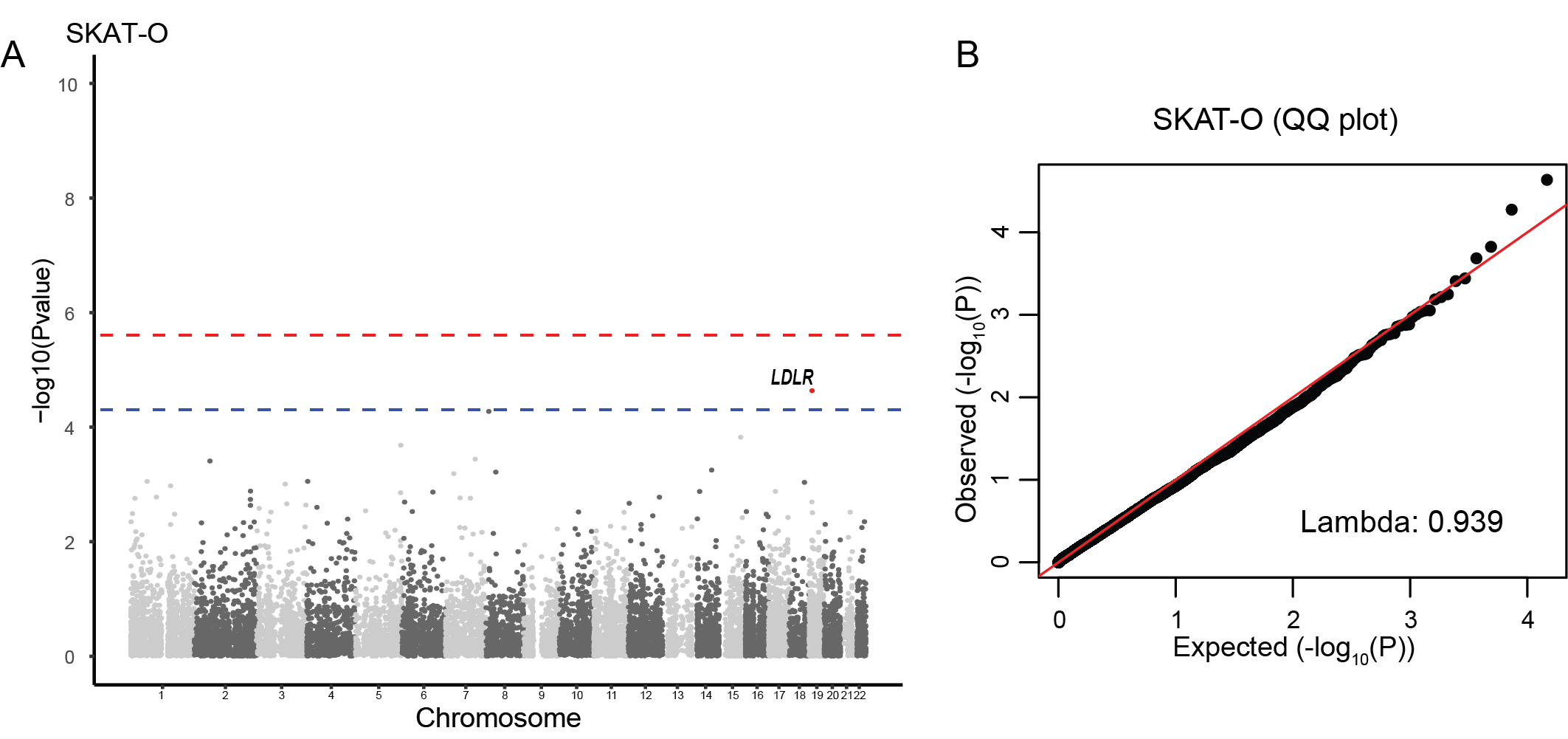

Manhattan plot (A) and QQ plot (B) of the aggregated rare variant association analysis for CAD in the discovery cohort of 1752 cases and 3019 controls using SAIGE-GENE+ software. The red and blue dashed line represents a gene-wide significance threshold of p=2.5*10^-6^ and a suggestive significance of p=5*10^-5^ threshold, respectively.

**Figure S5. Cross-validation framework in the machine learning-based analysis**

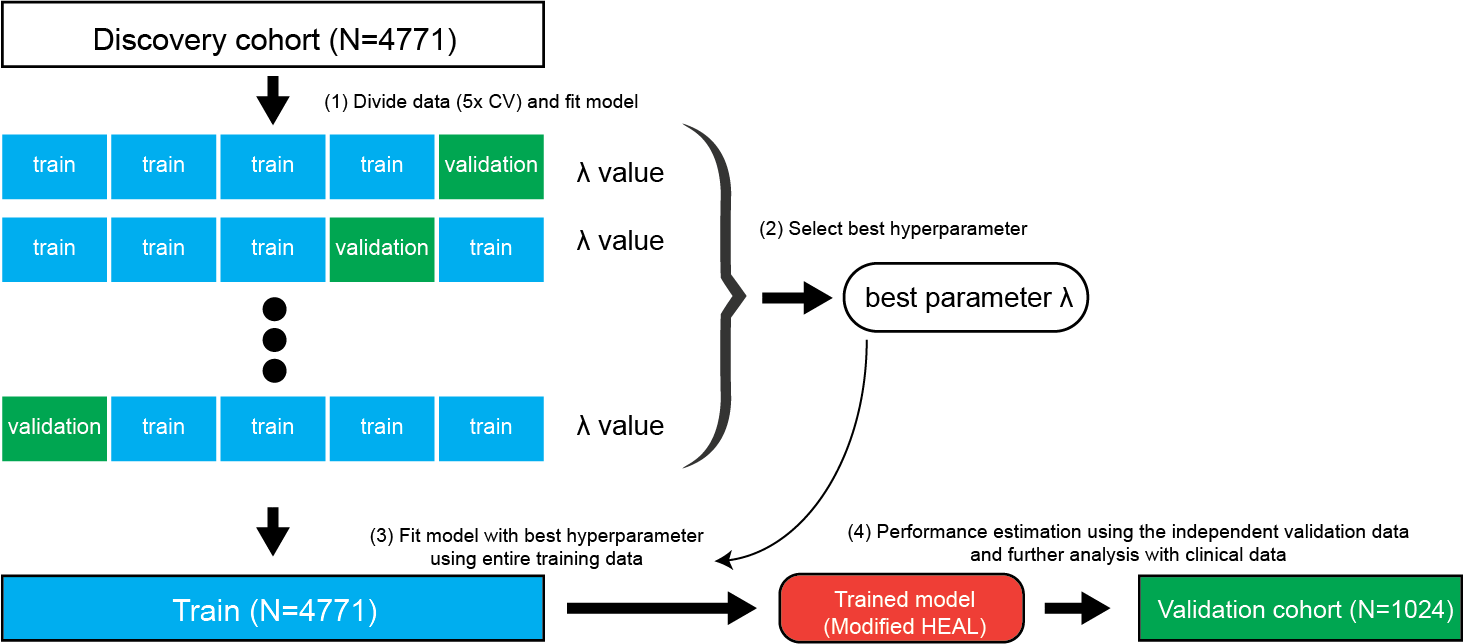

The discovery cohort (N = 4771) was divided into training and validation datasets for 5-fold cross-validation (CV) to determine the optimal hyperparameter (λ). For each fold, the data was split into training (blue) and validation (green) subsets and the model was fitted to the training data to evaluate different λ values. The best hyperparameter was selected based on performance across the folds. The model was then re-trained using the entire discovery cohort (N=4771) with the best hyperparameter. The trained model's performance was then estimated using an independent validation cohort (N=1024) to ensure robustness and further analyzed with clinical data.

**Figure S6. Cross-validation framework in the machine learning-based analysis**

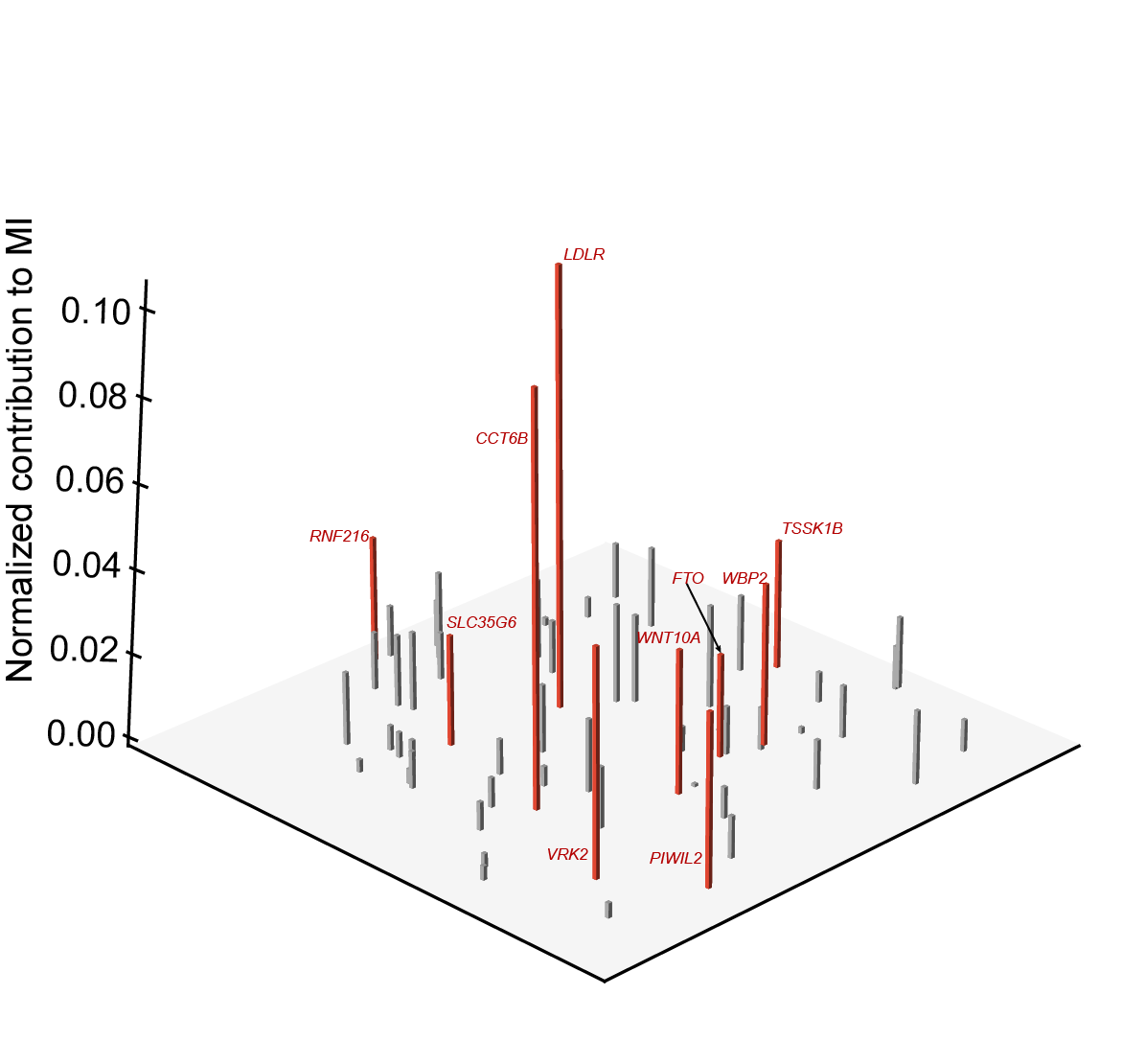

The mutational spectrum of CAD identified by the modified HEAL, in which 59 HEAL_CAD_ genes selected were randomly dispersed on the x-y plane. The bar heights (z-axis) for individual genes represent the corresponding contributions to CAD development, i.e., the coefficients learned by the modified HEAL. The top 10 disease-contributing genes are highlighted in red.

**Figure S7. Correlation between PRS and continuous clinical indices.**

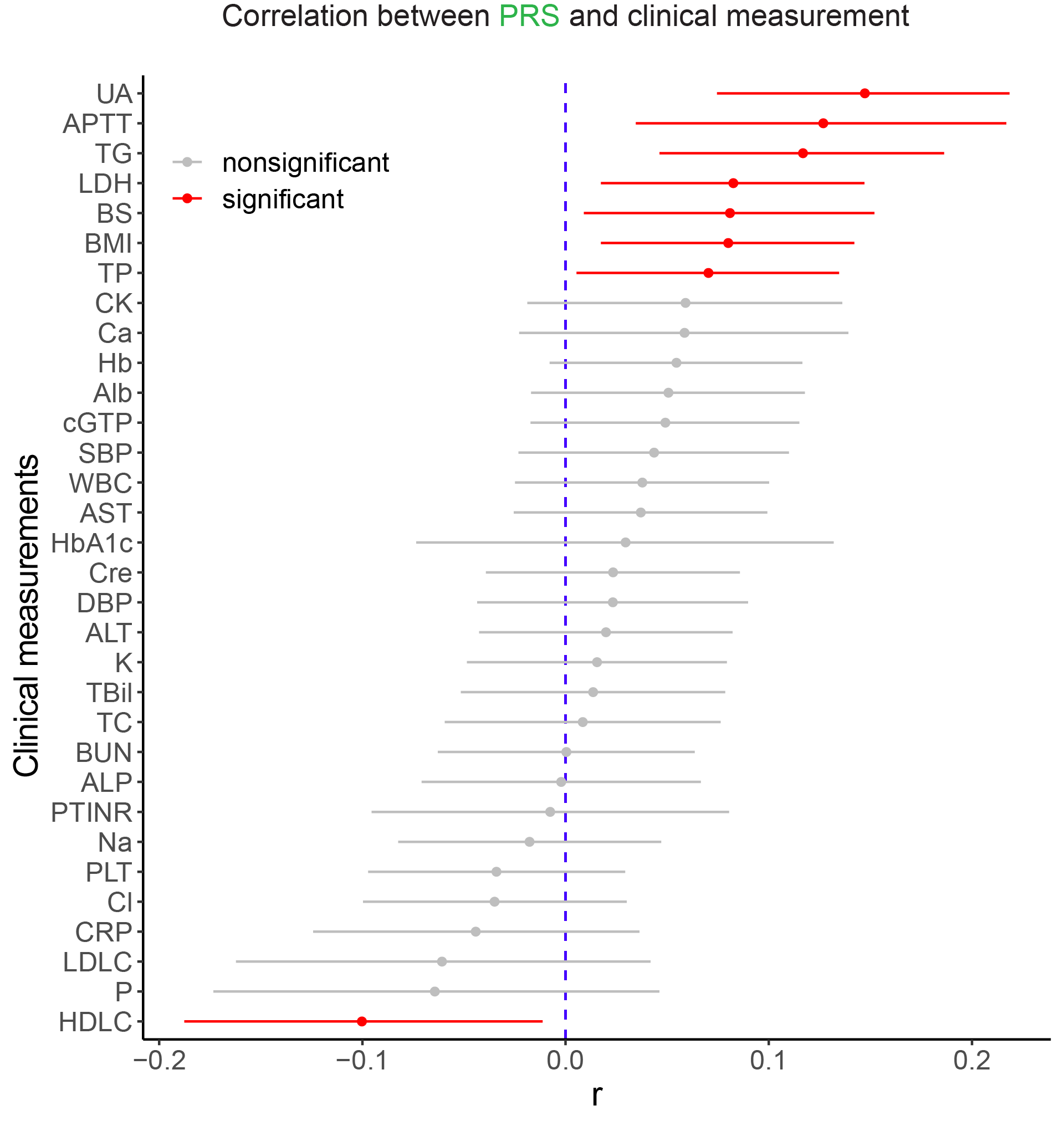
Data are presented as Pearson’s correlation coefficients and their 95% confidence intervals (CIs). Exact P values are shown in Table S10.

**Figure S8. Correlation between PRS and continuous clinical indices.**

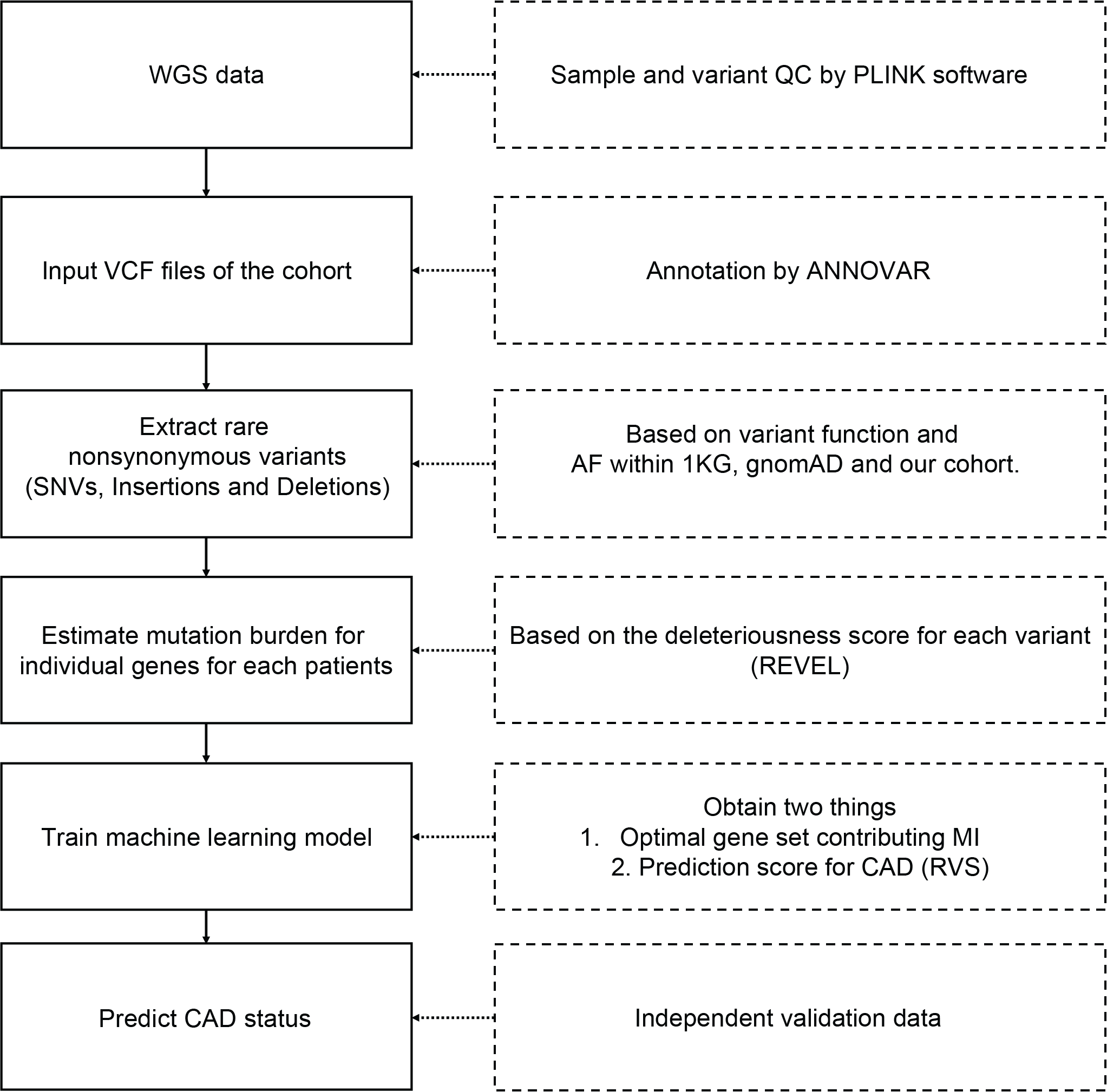

WGS, whole genome sequencing; QC, quality control; SNV, single nucleotide variant; AF, allele frequency; 1KG, 1000 Genomes Project dataset; CAD, coronary artery disease; RVS, rare variant-based risk score
